# Low-Intensity Focused Ultrasound of the Amygdala in Depression and Anxiety: A First-in-Human Active-Controlled Trial

**DOI:** 10.64898/2026.09.18.26363360

**Authors:** Amanda R. Arulpragasam, Mascha van ’t Wout-Frank, Yosef A. Berlow, Emily Aiken, Alison Gorbatov, Ryan Van Patten, Julia G. Gillotti, Hannah R. Swearingen, Christiana R. Faucher, Hannah Adams, Nicole C.R. McLaughlin, Israel Liberzon, Jennifer Barredo, Stephen Correia, Benjamin Greenberg, Noah S. Philip

## Abstract

**Background:** Low-intensity focused ultrasound (FUS) offers a noninvasive method for directly modulating deep brain structures with millimeter-scale precision, potentially addressing a major limitation of existing noninvasive neuromodulation approaches. The amygdala is a key node in affective neurocircuitry and a compelling target for psychiatric intervention. We conducted a first-in-human active controlled trial (NCT05147142) of amygdala-targeted FUS in patients with major depressive disorder, examining safety and target engagement.

**Methods:** Ten participants with major depressive disorder completed a single-blind randomized crossover trial with blinded symptom ratings, comparing imaging-guided FUS targeting the right amygdala versus left primary somatosensory cortex as an active control. FUS comprised two 10-minute applications within diagnostic ultrasound safety limits. Primary outcomes were safety and target engagement assessed with BOLD-fMRI during sonication, post-sonication arterial spin labeling (ASL) and resting-state functional connectivity. Secondary and exploratory outcomes included symptom change and imaging–symptom relationships.

**Results:** Nine participants completed both sessions. No serious adverse events occurred; neurological, neuropsychological, and MRI safety assessments were unremarkable. Adverse events were more frequent after amygdala than control sonication (p=0.028); one participant experienced clinical worsening following amygdala sonication and required monitoring. Amygdala sonication produced greater perfusion change in the targeted amygdala compared to the control region (p<0.001), and ipsilateral hippocampus (p<0.001). BOLD-fMRI during sonication demonstrated engagement of ventromedial prefrontal and rostral anterior cingulate regions, while post-sonication resting-state connectivity decreased between basolateral amygdala and sensorimotor regions (corrected *ps*<0.05). Symptom improvement did not differ between conditions, but exploratory imaging–symptom relationships were significant. Spontaneous reports of calm, clarity, or lightness occurred after amygdala but not control sonication (*p*<0.001).

**Conclusions:** Amygdala-targeted FUS produced anatomically specific, multimodal evidence of neuromodulation relative to active control, with a manageable safety profile. Convergent target and circuit-level signals support further development of FUS as a precision approach for modulating deep brain targets in psychiatric disorders.

## INTRODUCTION

Major depressive disorder (MDD) is a leading cause of disability worldwide and a substantial proportion of patients fail to achieve remission with first line treatments (1). This unmet need has driven interest in interventions that directly target dysfunctional neural circuits implicated in MDD (2). Neuromodulation has emerged as an important strategy within this framework. Deep brain stimulation demonstrates that circuit manipulation can produce clinical benefit (3), but its invasiveness limits scalability. Noninvasive approaches such as transcranial magnetic and electrical stimulation (4, 5), are more broadly adopted but cannot directly engage deep structures such as the amygdala, anterior cingulate, nucleus accumbens, and thalamus.

Low-intensity focused ultrasound (FUS) has emerged as a potentially transformative neuromodulation technology that combines noninvasiveness, reversibility, access to deep brain structures, and high spatial precision (reviewed in (6, 7)). Unlike high-intensity focused ultrasound, which produces thermal ablation (8), FUS operates within nonablative ranges intended to induce reversible physiological effects. These properties have generated substantial interest in FUS as a potential form of noninvasive deep brain stimulation and as a tool for causal interrogation of human neurocircuitry.

The amygdala is a compelling translational target, central to emotional learning, threat detection, salience attribution, autonomic regulation, and affective behavior (9, 10), with altered function implicated across a range of psychiatric disorders, including MDD, generalized anxiety disorder, posttraumatic stress disorder, schizophrenia, and substance use disorders. Its deep location and central role within frontolimbic networks (reviewed in (11, 12)) make the amygdala an attractive target for evaluating technologies capable of noninvasive deep brain modulation. Although the amygdala can be modulated indirectly (13, 14), direct, reversible perturbation in humans represents an important scientific opportunity. Recent reports indicate amygdala FUS is feasible in healthy individuals (15).

Although original work on FUS dates to the 1950s (16–18), recent preclinical and human work demonstrates that FUS can influence neural excitability (19, 20), neurovascular responses (21, 22), and synaptic plasticity (23, 24). In healthy humans, cortical and subcortical sonication can modulate cerebral perfusion (25, 26), resting-state functional connectivity (25), pain thresholds (27), and reward processing (28, 29). Emerging clinical studies targeting the amygdala (30), subcallosal cingulate cortex (31, 32), and nucleus accumbens (33), support the feasibility of noninvasive circuit modulation with apparently persistent effects. However, mechanisms and optimal stimulation parameters remain incompletely understood.

This uncertainty underscores the need for rigorous safety and mechanistic evaluation. While studies conducted within accepted safety limits generally report mild, transient effects (34), reports of brain injury following exposures outside these limits (35–38) and psychiatric worsening in a prior FUS study in MDD (31) highlight the need for careful safety monitoring, standardized exposure reporting, and direct assessment of target engagement (39, 40).

A fundamental translational challenge remains: can low-intensity FUS reliably and selectively engage an intended deep brain target in humans with sufficient precision to support mechanism-based psychiatric interventions? Thus, establishing target engagement is a prerequisite for efficacy testing. We reasoned that an essential first step is to demonstrate that acoustic energy can be delivered safely to a predefined deep limbic target, produce measurable biological effects within that region, and do so with anatomical specificity relative to an active control. In this framework, precision precedes efficacy: before asking whether FUS improves symptoms, it is necessary to determine whether it can reliably engage the neural circuitry it is intended to modulate.

We therefore conducted a first-in-human, randomized, active-controlled crossover study of MRI-guided FUS targeting the right amygdala in patients with MDD, with left primary somatosensory cortex (S1) serving as an active control. We hypothesized that FUS could be delivered safely within established exposure limits and produce evidence of target engagement. Clinical outcomes were exploratory and intended to inform future efficacy-focused investigations.

## METHODS

### Participants

Fifteen U.S. Veterans with MDD, with or without comorbid anxiety symptoms, provided written informed consent. Participants were recruited from the VA Providence Healthcare System. Diagnosis of MDD was verified using the Quick Structured Clinical Interview for DSM-5 (Quick-SCID). Eligible participants were 22–75 years of age, of any sex, and met DSM-5 criteria for MDD with clinically significant depressive symptoms based on standardized symptom rating scales. Participants receiving ongoing treatments were required to have stable treatment for at least 6 weeks. Women of childbearing potential were required to use an acceptable method of contraception and have a negative pregnancy test before FUS procedures.

Key exclusion criteria included contraindications to FUS or MRI, including a history of seizure disorder or serious neurological illness, structural or neurological abnormalities near the sonication site, prior brain surgery, implanted pacemakers or central nervous system devices, moderate or greater traumatic brain injury, recent head injury, greater than moderate alcohol or substance use disorder(s), metal in the head, or other conditions that could interfere with study assessments. Participants were also excluded for inability to comply with study procedures, acute suicidality, very severe symptom severity, or significant recent suicidal behavior as defined by the Columbia Suicide Severity Rating Scale (C-SSRS) and investigator assessment. The study was determined to be Significant Risk by the US Food and Drug Administration and thus conducted under Investigational Device Exemption G200146, approved by the VA Providence Institutional Review Board, and registered at ClinicalTrials.gov (NCT05147142).

### Study Design

Participants received two in-scanner FUS sessions in a randomized, single-blind crossover design, with one session targeting the right amygdala and the other targeting the left primary somatosensory cortex (S1) as an active control. FUS sessions were separated by at least one week. Participants were blinded to sonication condition, and MADRS assessments were performed by a psychiatrist blinded to amygdala or S1 target sonication. Prespecified safety outcomes included clinical MRI, neurological assessment, and neuropsychological testing, and were assessed at baseline, immediately following, and at 24-hours and 1-week after each FUS session (see **Safety Assessment** and **Figure 1**). Prespecified imaging outcomes included changes in cerebral perfusion, BOLD signal, and resting-state functional connectivity. Clinical symptom measures were evaluated as exploratory outcomes.

**Figure 1.**
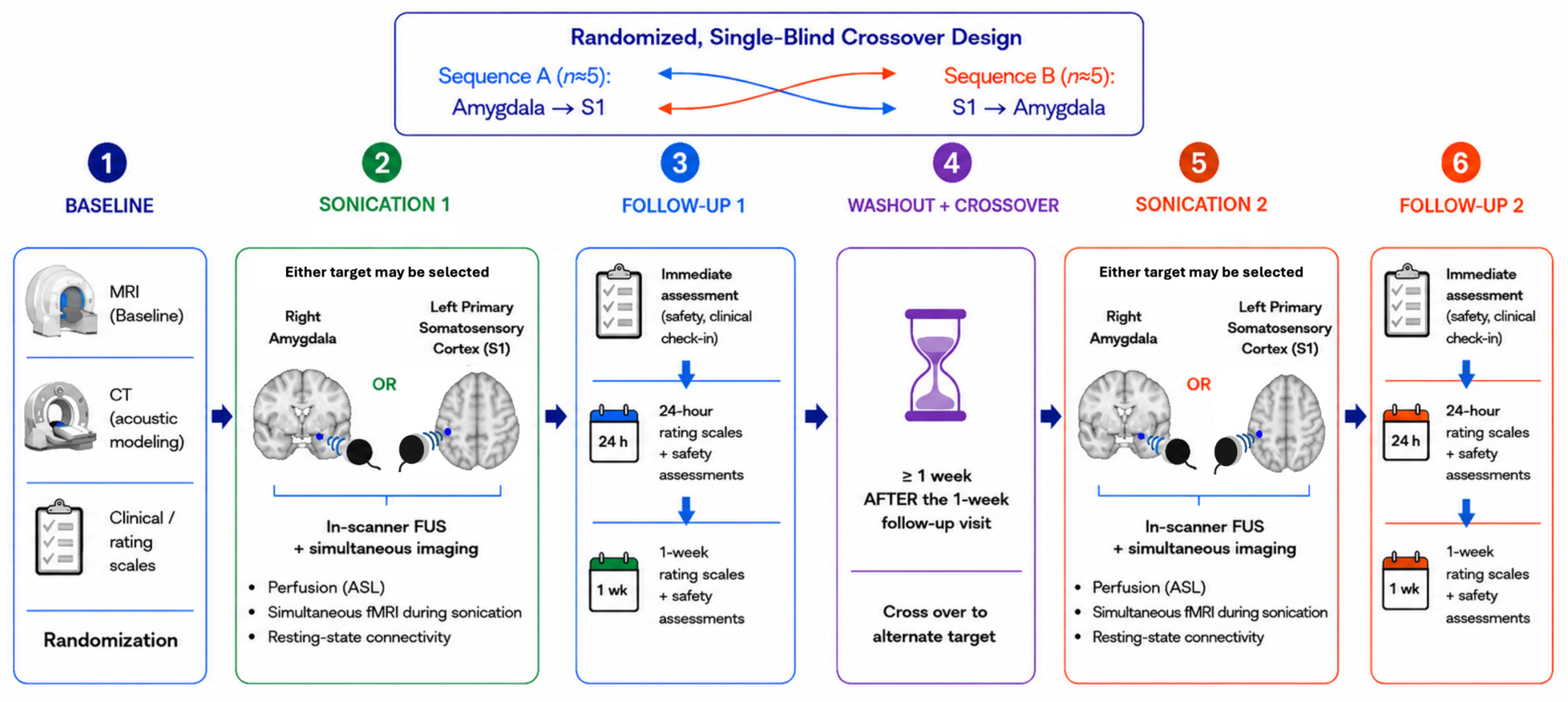
Study Design Overview. <u>Key:</u> ASL, Arterial spin labeling <u>Figure Legend.</u> Participants completed two randomized, single-blind in-scanner low-intensity focused ultrasound (FUS) sessions, with one session targeting the right amygdala and the other targeting the left primary somatosensory cortex (S1) as an active control. FUS sessions were separated by at least one week. Safety assessments were conducted at baseline, immediately following each FUS session, and at 24 hours and 1-week post-sonication. Imaging outcomes included cerebral perfusion, BOLD signal during FUS, and resting-state functional connectivity.

### FUS Protocol and Procedure

FUS was delivered using the Brainsonix Pulsar 1002 (Brainsonix, CA), an MRI-compatible, single-element 65mm transducer (fundamental frequency 650kHz; PRF 10Hz; pulse-width 5ms; hydrophone-measured peak focal depth 60.0mm, -6dB edges at 46.9mm and 77.0mm, 4mm focal width). Each session included two applications of ten sonications (30s on/off) separated by a brief break (<5min). All FUS was delivered inside of the MRI bore.

Device output was set at a Spatial-Peak Temporal Intensity (I_SPTA_) of 937.9 mW/cm2, and Spatial-Peak Pulse-Average Intensity (I_SPPA_) of 18.84 W/cm2, derated (0.3dB/cm/MHz) to intensities of _ISPTA.3_=719.5mW/cm^2^ and _ISPPA.3_=14.39 W/cm^2^ (peak negative pressure of 0.61 MPa and a mechanical index of 0.76). An investigator used scout/T1 MRI and device-specific fiducials for line-of-sight targeting of the amygdala, defined per the Mai Atlas (41), with placement verified by a second investigator. Coupling was performed using a 5-degree gel pad; commercial ultrasound gel was applied to the transducer face and inspected to ensure no bubbles were present; gel was also applied to the scalp, and the hair was manually combed in a single direction; the transducer was affixed to the skull via straps and secured within the MRI head coil (for further device information, see **Supplemental**).

### Safety Assessment

FUS safety was evaluated at baseline and repeated immediately, at 24-hours, and 1-week following FUS, interpreted relative to baseline. Adverse events were rated using the Systematic Assessment for Treatment Emergent Effects (SAFTEE; 96 symptoms across 17 domains), with additional spontaneous reports coded using the Medical Dictionary for Regulatory Activities (MedDRA); AEs following amygdala and S1 sonication were coded as separate episodes to permit comparison. Participants received clinical brain MRIs (T1, T2, SWI) with a stat radiologist read for FUS-induced injury, neurological examinations, and assessment with the C-SSRS.

Cognitive safety was assessed at baseline, at 24-hours, and 1-week using the Repeatable Battery for the Assessment of Neuropsychological Status (RBANS), Color Trails Trials (CTT), and the Neuropsychological Assessment Battery (NAB) Mazes test. This battery evaluated attention, processing speed, language, visuospatial skills, episodic memory, planning/organization, and cognitive flexibility, with a neuropsychologist reviewing scores for potential worsening. Alternate forms were used, with form assignment prespecified and balanced across participants to mitigate potential practice effects (see **Supplemental**).

### Computed Tomography

Participants underwent baseline Siemens head CT (0.45-mm in-plane resolution, 0.63-mm slice thickness, 120 kVp, 170 mA, BONEPLUS kernel) for subject-specific post hoc acoustic modeling.

### MRI Data Acquisition and Quality Control

Neuroimaging was acquired on a Siemens 3T Prisma. Because the transducer did not fit inside the 64-channel phased-array head coil, a 20-channel coil was used for simultaneous FUS-fMRI, with participants switched to the 64-channel coil for subsequent acquisitions. Functional images were collected during each FUS administration (20-channel; voxel size 2.5mm^3^, TR=700ms; TE=33.0ms, 54 slices; flip angle (FA)=70°; 10-minute acquisition). Resting state MRI was acquired at baseline and immediately following FUS (voxel size 2.5mm^3^, TR=700ms; TE=33.0ms, 54 slices; FA=70°; 12-minute acquisition). Arterial spin labeling (ASL) MRI was acquired at baseline and immediately following FUS (TR=4600ms, TE=16.18ms, 8 sequential slices, voxel size 1mm^3^, inversion time=1990ms, tag-controlled pulsed ASL (pASL), bolus duration=700ms; 5-minute acquisition). High resolution structural volumes were also collected at all imaging sessions (voxel size 1mm^3^, TR=2300ms; TE=2.98ms, FA=9°). Structural and functional MRI data were assessed using MRIQC in conjunction with visual inspection for artifacts; no participants or scans were excluded based on quality assessment (see **Supplemental**).

### MRI Data Analysis

*fMRI.* Functional images were preprocessed using fMRIPrep 24.1.1. Major steps include: 1) realignment, 2) slice time correction, 3) registration to MNI-152 volumetric and FreeSurfer spaces, and 4) spatial smoothing with a 5 mm full-width half-max (FWHM) Gaussian kernel using SPM25 (see **Supplemental**).

Change in fMRI BOLD signal during FUS delivery (10 × 30-sec long blocks of sonication separated by 30-sec rest blocks), was modeled as a single boxcar regressor convolved with the hemodynamic response function (HRF) in SPM25. Resting-state functional connectivity analyses were performed using the CONN toolbox (22.v2407). Seed-to-voxel analyses were performed with the basolateral amygdala (BLA) as an *a priori* seed (see **Supplemental**).

*Arterial Spin Labeling.* ASL images were preprocessed using the FSL-BASIL toolbox (standard preprocessing plus motion and partial volume correction). Perfusion-weighted images were normalized to participant-level mean grey matter perfusion to facilitate individual comparisons. Amygdala regions of interest (ROI) were functionally defined for each participant (see **Supplemental**). Individual subject cerebral blood flow (CBF) maps were registered to standard MNI152 space and analyzed at the group level using FSL’s randomise function with a voxel-wise GLM and 5,000 permutations. Threshold-Free Cluster Enhancement (TFCE) was applied, with family-wise error (FWE)-corrected *p*<0.05 considered significant.

### Rating Scales and Spontaneous Subjective Reports

Psychiatric symptoms were assessed at baseline, immediately following FUS, 24-hours post-FUS, and 1-week post-FUS using the Montgomery–Åsberg Depression Rating Scale (MADRS) (double-blind), Inventory of Depressive Symptomatology–Self-Report (IDS-SR), Generalized Anxiety Disorder-7 (GAD-7), PTSD Checklist for DSM-5 (PCL-5), and Clinical Global Impression (CGI; severity and improvement) scales. All participants were also asked, “What did you make of this?” within approximately 15 minutes following sonication. Spontaneous verbal responses were recorded for subsequent analysis.

### Statistical Analysis

Paired-sample *t*-tests tested whether perfusion changes were specific to target sonication. Symptom change over time (24 hours and 1 week) was analyzed using linear mixed models accounting for within-subject correlation, with sonication order included as a covariate. Associations between neuroimaging measures and symptom change were examined using correlation analyses. Hedges’ *g* is reported to adjust for small-sample bias. Statistical analyses were conducted using IBM SPSS Statistics, version 31.0. Statistical tests were two-sided, with *p* < 0.05 considered statistically significant. No additional multiple-comparison correction was applied to these analyses.

### Post Hoc Acoustic modeling

Post hoc subject-specific acoustic modeling was performed using BabelBrain (V0.4.2) with CT scans and planned targeting parameters at 650 kHz, estimating peak intensity and thermal effects and comparing simulated acoustic fields with the planned line-of-sight target to assess spatial overlap.

## RESULTS

**Table 1** summarizes participant demographics and clinical characteristics. The 10 participants ranged in age from 33 to 69 years (mean=47, SD=13); 8 (80%) were male and 2 (20%) were female. Participant flow is presented in the CONSORT diagram (**Figure 2**)

**Figure 2.**
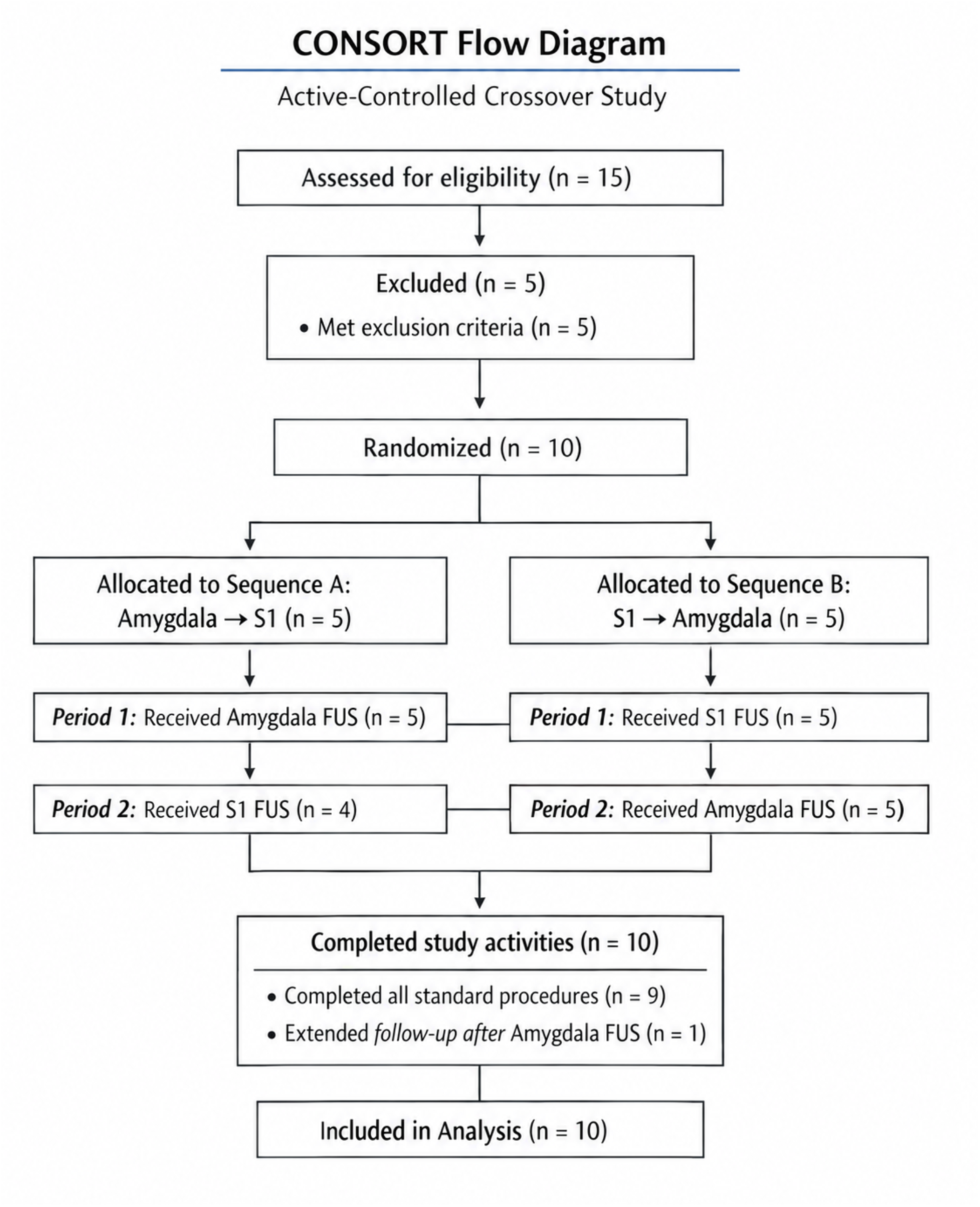
CONSORT Participant Flow Diagram. <u>Key:</u> FUS, low intensity focused ultrasound; S1, primary somatosensory cortex <u>Figure Legend.</u> Participant flow from assessment for eligibility through exclusion, randomization, allocation to FUS sequence, treatments received, and completion of study activities.

**Table 1.** Demographics.

| *Variables* | Participants (N=10) |  |
| --- | --- | --- |
|  | **Mean** | **SD** |
| Age (years) | 47 | 13 |
|  | ***n*** | ***%*** |
| Sex (female) | 2 | 20 |
| Race <sup>a</sup> |  |  |
| White | 8 | 80 |
| Asian | 1 | 10 |
| Multiracial | 1 | 10 |
| Prefer not to answer | 1 | 10 |
| Ethnicity <sup>a</sup> |  |  |
| Hispanic origin | 1 | 10 |
| Not of Hispanic origin | 7 | 70 |
| Prefer not to answer | 2 | 20 |
| Education <sup>a</sup> |  |  |
| Some college | 5 | 50 |
| Bachelor's degree | 2 | 20 |
| Some post-graduate education | 1 | 10 |
| Master's degree | 2 | 20 |
| Employment status <sup>a</sup> |  |  |
| Full time | 6 | 60 |
| Retired | 1 | 10 |
| Disabled | 3 | 30 |
| Service-connected disability (mental health) | 8 | 80 |
|  | ***n*** | ***%*** |
| *Psychiatric Comorbidities <sup>b</sup>* |  |  |
| Posttraumatic Stress Disorder | 6 | 60 |
| Generalized Anxiety Disorder | 5 | 50 |
| Panic Disorder | 2 | 20 |
| Attention Deficit Hyperactivity Disorder | 2 | 20 |
| Obsessive Compulsive Disorder | 1 | 10 |
| Bipolar II Disorder | 1 | 10 |
| Alcohol Use Disorder, Moderate | 1 | 10 |
| Cannabis Use Disorder, Mild | 1 | 10 |
| Binge Eating Disorder | 1 | 10 |
| Psychiatric History |  |  |
| Suicide attempt(s) | 2 | 20 |
| Inpatient hospitalization(s) | 2 | 20 |

Key: SD, Standard deviation
<sup>a</sup> Totals may not equal 100% due to non-response or multiple responses; demographic variables were assessed via self-report; <sup>b</sup> comorbidities based on structured clinical interview and chart review.

### Safety and Tolerability

Assessments indicated predominantly mild transient AEs with no serious adverse events, unanticipated device events, evidence of brain injury or sustained cognitive decline. The most common AEs were headache, fatigue, and sleep problems following both amygdala and S1 sonication, though more AEs occurred after amygdala sonication (2.50 vs 1.11 events/exposure; RR, 2.25; 95% CI, 1.08–4.68; exact Poisson p=0.028) (**Table 2**). No MRI-detectable brain injury was identified across 61 post-FUS clinical scans, and no FUS-related neurological abnormalities or worsening of suicidality were observed. Neuropsychological function remained largely unchanged, with nine participants’ scores within ±2 SD of baseline (see **Supplemental**); one participant improved by 2.58 SD on the RBANS Attention Index at 1-week post-amygdala sonication.

**Table 2.** Adverse Events.

| **Adverse Event Category** | **Amygdala**<br>**(n=10)** | **S1 (n=9)** |
| --- | --- | --- |
| **Mood** | 1 (10%) | 0 (0%) |
| **Cognitive** | 3 (30%) | 1 (11.1%) |
| **Visual Changes** | 2 (20%) | 0 (0%) |
| **Fatigue** | 3 (30%) | 3 (3.33%) |
| **Headache** | 4 (40%) | 3 (3.33%) |
| **Sonication site discomfort** | 1 (10%) | 0 (0%) |
| **Hyperhidrosis** | 1 (10%) | 0 (0%) |
| **Auditory** | 1 (10%) | 0 (0%) |
| **Sleep problems** | 3 (30%) | 2 (22.2%) |
| **Device discomfort** | 2 (20%) | 0 (0%) |
| **Hyperactivity** | 1 (10%) | 0 (0%) |
| **Other pain** | 0 (0%) | 1 (11.1%) |
| **Vegetative** | 2 (20%) | 0 (0%) |
| **Dissociation** | 1 (10%) | 0 (0%) |
| **Total** | 25 | 10 |

Key: S1, primary somatosensory cortex.

### Arterial Spin Labeling

Perfusion changes were significantly greater in the targeted right amygdala following FUS than in either the contralateral S1 control (t(9)=5.15, p<0.001; Hedges’ g=1.55) or the adjacent right hippocampal head (t(9)=4.70, p<0.001; Hedges’ g=1.36 (**Figure 3A**), whereas S1 FUS produced no significant change in left S1 perfusion (p>0.1). At the group level, right amygdala FUS was associated with significant perfusion increases in the bilateral amygdala and right rACC, surviving small-volume FWE correction (left amygdala pFWE<0.01, right amygdala pFWE=0.02, right rACC pFWE<0.01; **Figure 3B**). Individual trajectories similarly showed greater changes in ASL perfusion within the right amygdala following amygdala sonication, although the direction of change varied across participants (**Supplementary Figure S1A**), whereas perfusion changes in the control S1 region were unchanged following control sonication (**Supplementary Figure S1B**).

**Figure 3.**
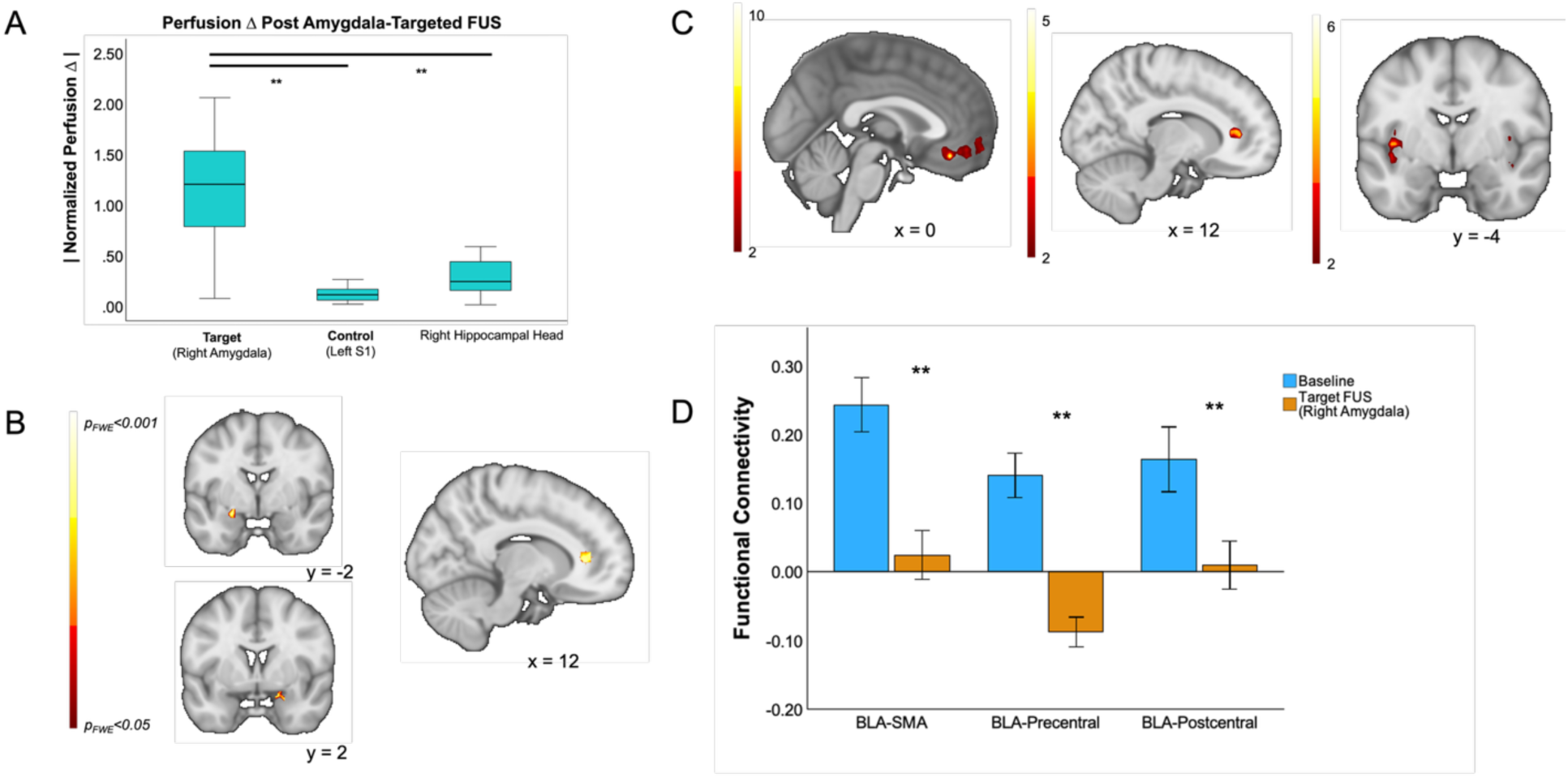
MRI Measures of Low-Intensity Focused Ultrasound-Induced Neural Effects. <u>Key:</u> MRI, Magnetic resonance imaging; FUS, low intensity focused ultrasound; S1, primary somatosensory cortex. ** indicates p<.001. <u>Figure Legend.</u> (A) Absolute change in perfusion following right amygdala FUS compared with control S1 FUS and the adjacent right hippocampal head. (B) Group-level ASL perfusion changes following right amygdala FUS, showing significant increases in the bilateral amygdala and right rostral anterior cingulate cortex (rACC) following small-volume correction. (C) Brain regions showing increased BOLD signal during FUS-on compared with FUS-off periods during right amygdala FUS, including the ventromedial prefrontal cortex (vmPFC), right rACC, and left insula. (D) Decreases in resting-state functional connectivity between the right basolateral amygdala (BLA) and the left supplementary motor area (SMA), right precentral gyrus, and left postcentral gyrus following right amygdala FUS compared with baseline.

### FUS-Induced BOLD Activation

During amygdala FUS, BOLD signal increased during FUS-on relative to FUS-off periods in the vmPFC (peak pFWE=0.006), right rACC (cluster pFWE=0.020), and left insula (cluster pFWE=0.042; **Figure 3C**), all surviving small-volume correction (see **Supplemental**). Because transcranial FUS can produce auditory sensations and auditory stimulation can contribute to online FUS effects (42–44), we examined primary auditory cortex activation. S1 FUS significantly activated right (x=57, y=-4, z=2; pFWE=0.018) and left (x=-44, y=-21, z=14; pFWE=0.002) auditory cortex, with no significant activation during amygdala FUS. Direct comparison showed significantly greater auditory cortex activation during S1 than amygdala FUS in both right (x=59, y=-9, z=2; pFWE=0.014) and left (x=-52, y=-21, z=10; pFWE=0.006) hemispheres (**Supplemental Figure S2**).

### Resting-State Functional Connectivity

Seed-to-voxel analyses using the right basolateral amygdala (BLA) as the *a priori* seed identified significant decreases in resting-state functional connectivity (rsFC) following amygdala FUS compared with baseline. Decreased rsFC was observed between the right BLA and the left postcentral gyrus, right precentral gyrus, and left supplementary motor area (SMA; *p*FDR<0.05). Compared with the control sonication site, amygdala FUS was also associated with decreased rsFC between the right BLA and the right precentral and postcentral gyri (*p*FDR<0.05). No significant changes in right BLA rsFC were observed following control sonication compared with baseline (*p*FDR>0.05; **Figure 3D**). Sensitivity analyses accounting for between-session changes in head motion yielded similar decreases in all three findings (see **Supplemental**).

### Psychiatric Symptoms and Spontaneous Reports

Linear mixed-effects models showed significant main effects of time across all symptom measures (MADRS: F(4, 30.17)=10.64, p<0.001; IDS-SR: F(4, 30.21)=5.90, p=0.001; PCL-5: F(4, 30.09)=10.31, p<0.001; GAD-7: F(4, 30.17)=3.58, p=0.02; CGI-S: F(6, 45.30)=5.36, p<0.001; CGI-I: F(5, 36.05)=3.14, p=0.019), indicating improvement over time (**Figure 4A**). Neither sonication order nor the time×order interaction was significant for any measure except for a significant time×order interaction for CGI-I, *F*(5, 36.05)=4.48, *p*=.003). Following amygdala-targeted FUS, 7/10 participants spontaneously described increased clarity, lightness, or calm; no participants (0/10) reported these experiences following S1 FUS (p<.001).

**Figure 4.**
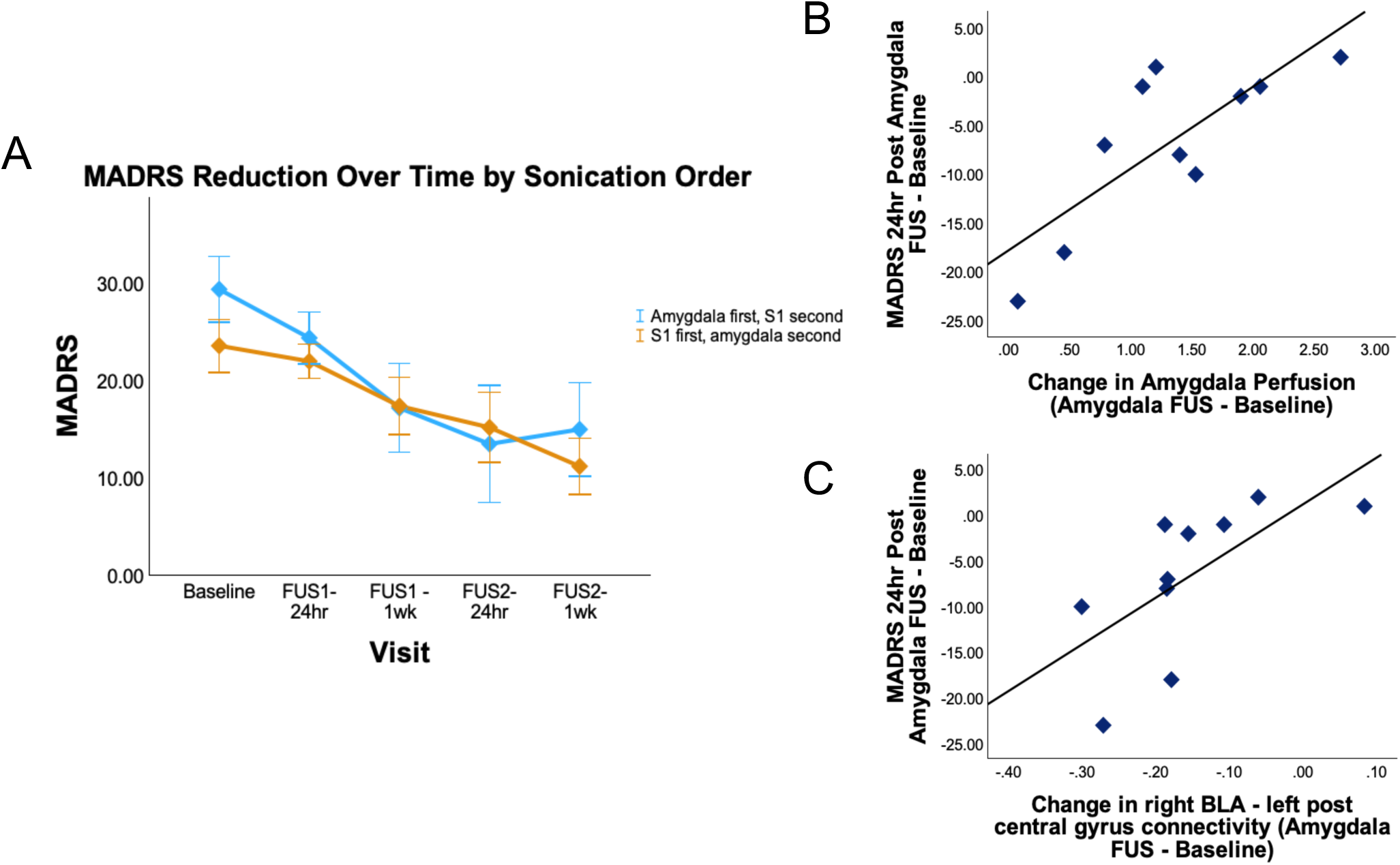
Psychiatric Symptom Trajectories and Associations with Imaging Measures. <u>Key:</u> FUS, low intensity focused ultrasound; MADRS, Montgomery-Åsberg Depression Rating Scale (double-blind); S1, primary somatosensory cortex. BLA, basolateral amygdala. <u>Figure Legend.</u> (A) Changes in MADRS scores over time, shown separately by sonication order. (B) Association between change in right amygdala perfusion and change in MADRS score 24 hours following amygdala-targeted FUS. (C) Association between change in resting-state functional connectivity between the right basolateral amygdala (BLA) and left postcentral gyrus and change in MADRS score 24 hours following amygdala-targeted FUS.

### Associations between Psychiatric Symptoms and Imaging Measures

We examined associations between changes in imaging measures and psychiatric symptoms at 24-hours and 1-week following sonication, Changes in amygdala perfusion were significantly associated with changes in MADRS scores following amygdala FUS at 24 hours, with smaller perfusion changes associated with greater depression symptom improvement (*r*=0.787, *p*=0.007; **Figure 4B**) and 1-week (*r*=0.660, *p*=0.038). No significant correlations were observed between changes in S1 perfusion and changes in psychiatric symptoms 24-hours following S1 sonication (all *p*s>0.05). Changes in right BLA connectivity with the left postcentral gyrus were also significantly associated with changes in MADRS scores at 24-hours, such that greater decreases in connectivity were associated with greater reductions in depressive symptoms (*r*=0.729, *p*=0.017; **Figure 4C**). At 1-week, this relationship was not significant. Additional exploratory associations between changes in imaging measures and IDSSR, GAD-7, PCL-5, and CGI scores are reported in the **Supplement.**

### Clinical Course Following Amygdala FUS (n=1)

One participant demonstrated protocol-defined clinically significant worsening in depressive symptoms 24-hours following amygdala-targeted FUS, that remained elevated at 1-week; this triggered the a priori safety monitoring plan that included cessation of further FUS and weekly assessments for one month (see **Supplement**). This participant also showed a marked perfusion increase in the targeted region (441% above baseline), with modeled acoustic pressure positively associated with regional perfusion (r=0.26, p=0.007). Both perfusion and depressive symptoms returned to near-baseline within the 1-month follow-up (**Supplemental Figure S3**).

### Acoustic Modeling and Targeting Accuracy

Mean displacement between modeled and line-of-sight target locations was 1.75±0.87mm (x), - 0.46±0.64mm (y), and -4.81±2.80mm (z), with the greatest deviation along the z-axis; overall, modeled beam locations showed relatively small deviations from line-of-sight targets (**Supplemental Figure S4A**). Notably, the participant who demonstrated clinical worsening had the greatest x-direction (lateral) deviation between line-of-sight targeting and the modeled beam alongside a change in beam morphology (**Supplemental Figure S4B**).

## DISCUSSION

This first-in-human active-controlled study in MDD provides convergent evidence that low-intensity focused ultrasound can selectively engage the amygdala and modulate distributed circuitry relevant to affective psychopathology. Amygdala-targeted sonication produced focal perfusion changes specific to the intended target relative to both an active control and the adjacent hippocampal head, while complementary neuroimaging measures demonstrated effects across interconnected regions. These findings provide proof-of-principle that FUS can noninvasively, and with anatomical specificity, modulate a deep limbic structure in patients with psychiatric illness, addressing a longstanding limitation of noninvasive neuromodulation.

Perfusion provided the strongest evidence for spatial selectivity. Post hoc subject-specific acoustic modeling demonstrated minimal deviations between modeled beam locations and planned line-of-sight targets, with the largest displacement along the z-axis (i.e., depth). These findings support, but do not definitively establish, targeting accuracy given the limitations of current methods for measuring acoustic fields within the intact human brain.

Amygdala sonication also produced effects beyond the focal target. Group-level perfusion increased in the bilateral amygdala and ipsilateral rACC, while FUS-on versus FUS-off BOLD responses involved the vmPFC, rACC, and insula (comparable to prior findings in older healthy controls (25)) and resting-state analyses identified subsequent changes in basolateral amygdala connectivity with sensorimotor regions. These findings complement prior work demonstrating acute amygdala BOLD modulation with FUS in patients with mood, anxiety, and trauma-related disorders (28). In that study, active relative to sham FUS also affected the adjacent hippocampus; in the present study, perfusion change was substantially greater in the targeted amygdala than in the hippocampal head, providing complementary evidence for anatomical selectivity. Together, these multimodal findings indicate that focal amygdala sonication can produce measurable local and distributed circuit effects.

Although this study was not designed or powered to evaluate clinical efficacy, psychiatric symptoms improved over time across multiple measures. The absence of an order effect, together with the crossover design and the possibility of prolonged biological effects, limits attribution of these changes to amygdala FUS. Nevertheless, smaller changes in amygdala perfusion were associated with greater MADRS improvement at 24 hours and 1-week, and greater decreases in right BLA–left postcentral gyrus connectivity were associated with greater MADRS improvement at 24 hours. Conversely, the participant with protocol-defined clinical worsening demonstrated the largest increase in amygdala perfusion. Taken together, the perfusion findings raise the possibility that the relationship between physiological target engagement and clinical effect may be nonlinear, such that greater perturbation is not necessarily associated with greater benefit. This hypothesis is consistent with the concept of an optimal biological response window and should be tested prospectively in dose-ranging studies. Spontaneous reports of increased calm, clarity, or lightness following amygdala FUS provide an additional indication of potential effects. Given the small sample, these findings are hypothesis-generating rather than evidence of efficacy and warrant evaluation in larger, adequately powered studies.

Safety findings were generally reassuring and consistent with prior studies (e.g., (30, 31)) but underscore the need for careful monitoring. Adverse events were predominantly mild and transient, with no SAEs or evidence of injury, although AEs were more frequent after amygdala sonication. One participant experienced protocol-defined worsening of depressive symptoms accompanied by a marked increase in amygdala perfusion; both resolved during follow-up. This finding illustrates that greater physiological robust target engagement should not be assumed to confer greater therapeutic benefit; consistent with prior work in healthy individuals showing that amygdala FUS increased arousal to negative images (25). The positive association between modeled acoustic pressure and regional perfusion in this participant cannot establish causality and reinforces the need for prospective dose-ranging studies integrating safety and biological target-engagement measures.

This study has several limitations, including its small, predominantly male and White sample, single-session design, and inability to establish dose-response relationships or efficacy. Because precision was prioritized over efficacy a full sham condition was not included; future studies should consider sham and anatomical active-control conditions. The crossover design may also have been susceptible to carryover effects if FUS effects persist beyond the treatment session, which would favor parallel-group designs or longer washout periods. Functional imaging was obtained at only a single post-sonication timepoint, reflecting the brief duration of macaque findings available during study design (45), thus limiting the characterization of potential temporal changes. In-scanner sonication enabled direct verification of transducer placement but limit generalizability to non-MRI-guided FUS. We also did not incorporate auditory masking, although the absence of auditory cortex activation during amygdala FUS provides some reassurance against a major auditory confound.

Current methods cannot directly measure the intracranial location of the applied beam, although acoustic radiation force imaging (46) may eventually enable direct assessment. Post hoc acoustic modeling supported correspondence between the modeled beam and planned target locations should be interpreted with caution. We also did not obtain other measures of amygdala output (e.g., pupillary response, heart rate). Lastly, we report both the absolute magnitude and direction of ASL perfusion change. Absolute change was emphasized as a measure of the magnitude of physiological perturbation, whereas directionality was interpreted cautiously because increases or decreases in ASL may reflect different combinations of vascular and anatomical factors.

Taken together, these findings demonstrate that low-intensity FUS can noninvasively engage the human amygdala with anatomically specific physiological effects that extend to distributed circuitry. The convergence of perfusion, network-level changes, and exploratory symptom associations supports further investigation of amygdala FUS as a means of noninvasively modulating deep neural circuits relevant to psychiatric illness, providing a foundation for larger, parallel-group and repeated-dose studies to define optimal dosing, durability, safety, and clinical benefit.

## Data Availability

All data produced in the present work are contained in the manuscript

## Use of AI statement

During paper drafting, ChatGPT (OpenAI, GPT-5.5) and Claude (Anthropic, Sonnet 5) were used to assist with editing for grammar, clarity, and concision, and to generate the first draft of the conceptual study design overview. The authors independently reviewed and revised all content and take full responsibility for the final manuscript.

## ACKNOWELDGEMENTS

The authors gratefully acknowledge Nathan McDannold PhD, Taylor Kuhn PhD, Natalie Rotstein BS, Miriam Goldberg MD PhD, and Steve Mernoff MD, for their contributions to study procedures, and all our participants for their time and trust.

## Clinical trials identifier

NCT05147142

## US Food and Drug Investigational Device Exemption

G200146

## References

1. Friedrich MJ. Depression Is the Leading Cause of Disability Around the World. JAMA. 2017;317(15):1517.

2. Siddiqi SH, Schaper FLWVJ, Horn A, Hsu J, Padmanabhan JL, Brodtmann A, et al. Brain stimulation and brain lesions converge on common causal circuits in neuropsychiatric disease. Nature Human Behaviour. 2021;5(12):1707–16.

3. Johnson KA, Okun MS, Scangos KW, Mayberg HS, de Hemptinne C. Deep brain stimulation for refractory major depressive disorder: a comprehensive review. Mol Psychiatry. 2024;29(4):1075–87.

4. Deng ZD, Lisanby SH, Peterchev AV. Electric field depth-focality tradeoff in transcranial magnetic stimulation: simulation comparison of 50 coil designs. Brain Stimul. 2013;6(1):1–13.

5. Philip NS, Nelson BG, Frohlich F, Lim KO, Widge AS, Carpenter LL. Low-Intensity Transcranial Current Stimulation in Psychiatry. American Journal of Psychiatry. 2017;174(7):628–39.

6. Arulpragasam AR, Theyel B, Barredo J, Pouille F, Gillotti JG, Greenberg BD, et al. Low-Intensity Focused Ultrasound Neuromodulation in Psychiatric Disorders: Mechanisms, Models, and Missing Links. Biol Psychiatry. 2026.

7. Davidson B, Xhima K, Cosgrove R, Hamani C, Eitan R, Rezai A, et al. A roadmap for focused ultrasound applications in psychiatry: Proceedings of the 2024 symposium on focused ultrasound in psychiatry (FUS-PULSE). Brain Stimul. 2025;18(5):1651–62.

8. De Schlichting E, Meng Y, Huang Y, Jones RM, Hynynen K, Hamani C, et al. Magnetic resonance-guided ultrasound thalamotomy for essential tremor: a review. Expert Rev Med Devices. 2025;22(9):989–97.

9. Fox AS, Shackman AJ. An Honest Reckoning With the Amygdala and Mental Illness. American Journal of Psychiatry. 2024;181(12):1059–75.

10. Grogans SE, Fox AS, Shackman AJ. The Amygdala and Depression: A Sober Reconsideration. American Journal of Psychiatry. 2022;179(7):454–7.

11. McTeague LM, Rosenberg BM, Lopez JW, Carreon DM, Huemer J, Jiang Y, et al. Identification of Common Neural Circuit Disruptions in Emotional Processing Across Psychiatric Disorders. Am J Psychiatry. 2020;177(5):411–21.

12. Ressler KJ. Amygdala activity, fear, and anxiety: modulation by stress. Biol Psychiatry. 2010;67(12):1117–9.

13. Sydnor VJ, Cieslak M, Duprat R, Deluisi J, Flounders MW, Long H, et al. Cortical-subcortical structural connections support transcranial magnetic stimulation engagement of the amygdala. Science Advances. 2022;8(25):eabn5803.

14. van Rooij SJH, Langhinrichsen-Rohling R, Minton ST, Hinojosa CA, Lukemire J, Lipschutz R, et al. Personalized fMRI-Guided TMS Targeting the Threat Neurocircuitry in PTSD: A Randomized Clinical Trial. American Journal of Psychiatry. 2026;183(5):343–54.

15. Algermissen J, Rascu M, Weber LA, den Boer T, Martin E, Treeby B, et al. Low-intensity focused ultrasound to human amygdala reveals a causal role in ambiguous emotion processing and alters local and network activity. Neuron. 2026;114(7):1269–89.e8.

16. Fry FJ, Ades HW, Fry WJ. Production of Reversible Changes in the Central Nervous System by Ultrasound. Science. 1958;127(3289):83–4.

17. Fry FJ, Barger JE. Acoustical properties of the human skull. The Journal of the Acoustical Society of America. 1978;63(5):1576–90.

18. Fry WJ, Wulff VJ, Tucker D, Fry FJ. Physical Factors Involved in Ultrasonically Induced Changes in Living Systems: I. Identification of Non-Temperature Effects. The Journal of the Acoustical Society of America. 1950;22(6):867–76.

19. Tufail Y, Matyushov A, Baldwin N, Tauchmann ML, Georges J, Yoshihiro A, et al. Transcranial pulsed ultrasound stimulates intact brain circuits. Neuron. 2010;66(5):681–94.

20. Tyler WJ, Tufail Y, Finsterwald M, Tauchmann ML, Olson EJ, Majestic C. Remote excitation of neuronal circuits using low-intensity, low-frequency ultrasound. PLoS One. 2008;3(10):e3511.

21. Iida K, Luo H, Hagisawa K, Akima T, Shah PK, Naqvi TZ, et al. Noninvasive low-frequency ultrasound energy causes vasodilation in humans. J Am Coll Cardiol. 2006;48(3):532–7.

22. Shen YY, Jethe JV, Reid AP, Hehir J, Amaral MM, Ren C, et al. Label free, capillary-scale blood flow mapping in vivo reveals that low-intensity focused ultrasound evokes persistent dilation in cortical microvasculature. Commun Biol. 2025;8(1):12.

23. Meng D, Zhang C, Pei J, Zhang X, Lu H, Ji H, et al. Low-intensity transcranial ultrasound stimulation promotes the extinction of fear memory through the BDNF-TrkB signaling pathway. Neuroimage. 2025;319:121441.

24. Pei J, Zhang C, Zhang X, Zhao Z, Zhang X, Yuan Y. Low-intensity transcranial ultrasound stimulation improves memory in vascular dementia by enhancing neuronal activity and promoting spine formation. Neuroimage. 2024;291:120584.

25. Hoang-Dang B, Halavi SE, Rotstein NM, Spivak NM, Dang NH, Cvijanovic L, et al. Transcranial Focused Ultrasound Targeting the Amygdala May Increase Psychophysiological and Subjective Negative Emotional Reactivity in Healthy Older Adults. Biol Psychiatry Glob Open Sci. 2024;4(5):100342.

26. Kuhn T, Spivak NM, Dang BH, Becerra S, Halavi SE, Rotstein N, et al. Transcranial focused ultrasound selectively increases perfusion and modulates functional connectivity of deep brain regions in humans. Front Neural Circuits. 2023;17:1120410.

27. Badran BW, Caulfield KA, Stomberg-Firestein S, Summers PM, Dowdle LT, Savoca M, et al. Sonication of the anterior thalamus with MRI-Guided transcranial focused ultrasound (tFUS) alters pain thresholds in healthy adults: A double-blind, sham-controlled study. Brain Stimul. 2020;13(6):1805–12.

28. Chou T, Kochanowski BJ, Hayden A, Borron BM, Barbeiro MC, Xu J, et al. A Low-Intensity Transcranial Focused Ultrasound Parameter Exploration Study of the Ventral Capsule/Ventral Striatum. Neuromodulation. 2025;28(1):146–54.

29. Yaakub SN, Eraifej J, Bault N, Lojkiewiez M, Bellec E, Roberts J, et al. Non-invasive ultrasonic neuromodulation of the human nucleus accumbens impacts reward sensitivity. Nat Commun. 2025;16(1):10192.

30. Barksdale BR, Enten L, DeMarco A, Kline R, Doss MK, Nemeroff CB, et al. Low-intensity transcranial focused ultrasound amygdala neuromodulation: a double-blind sham-controlled target engagement study and unblinded single-arm clinical trial. Molecular Psychiatry. 2025;30(10):4497–511.

31. Riis TS, Feldman DA, Kwon SS, Vonesh LC, Koppelmans V, Brown JR, et al. Noninvasive Modulation of the Subcallosal Cingulate and Depression With Focused Ultrasonic Waves. Biol Psychiatry. 2025;97(8):825–34.

32. Attali D, Tiennot T, Manuel TJ, Daniel M, Houdouin A, Annic P, et al. Deep transcranial ultrasound stimulation using personalized acoustic metamaterials improves treatment-resistant depression in humans. Brain Stimul. 2025;18(3):1004–14.

33. Mahoney Iii JJ, Thompson-Lake DGY, Johnson L, Ait-Daoud N, Lynch WJ, Olaitan G, et al. Focused Ultrasound Neuromodulation for Substance Use Disorder. Biological Psychiatry.

34. Legon W, Adams S, Bansal P, Patel PD, Hobbs L, Ai L, et al. A retrospective qualitative report of symptoms and safety from transcranial focused ultrasound for neuromodulation in humans. Scientific Reports. 2020;10(1):5573.

35. Rezai A, Ranjan M, Bhagwat A, Arsiwala T, Carpenter J, Schafer M, et al. Brain injury during focused ultrasound neuromodulation for substance use disorder. Brain Stimul. 2025;18(6):2050–3.

36. Rezai A, Ranjan M, Bhagwat A, Arsiwala T, Carpenter J, Schafer M, et al. Further clarification regarding brain injury during focused ultrasound neuromodulation for substance use disorder. Brain Stimul. 2026;19(1):103014.

37. Fouragnan E. Response to: Brain injury during focused ultrasound neuromodulation for substance use disorder. Brain Stimul. 2026;19(1):102989.

38. Klein-Flugge MC, Airan RD, Attali D, Aubry JF, Bubrick EJ, Caskey CF, et al. Open letter on intervention regimes and adverse events in focused ultrasound for neuromodulation. Brain Stimul. 2026;19(1):102994.

39. Aubry J-F, Attali D, Schafer ME, Fouragnan E, Caskey CF, Chen R, et al. ITRUSST consensus on biophysical safety for transcranial ultrasound stimulation. Brain Stimulation: Basic, Translational, and Clinical Research in Neuromodulation. 2025;18(6):1896–905.

40. Martin E, Aubry J-F, Schafer M, Verhagen L, Treeby B, Pauly KB. ITRUSST consensus on standardised reporting for transcranial ultrasound stimulation. Brain Stimulation: Basic, Translational, and Clinical Research in Neuromodulation. 2024;17(3):607–15.

41. Mai JK MM, Paxinos G. Atlas of the Human Brain. 4th ed: Academic Press; 2015 2015.

42. Guo H, Hamilton M, II, Offutt SJ, Gloeckner CD, Li T, Kim Y, et al. Ultrasound Produces Extensive Brain Activation via a Cochlear Pathway. Neuron. 2018;98(5):1020–30.e4.

43. Kop BR, Oghli YS, Grippe TC, Nandi T, Lefkes J, Meijer SW, et al. Auditory confounds can drive online effects of transcranial ultrasonic stimulation in humans. eLife Sciences Publications, Ltd; 2024.

44. Sato T, Shapiro MG, Tsao DY. Ultrasonic Neuromodulation Causes Widespread Cortical Activation via an Indirect Auditory Mechanism. Neuron. 2018;98(5):1031–41.e5.

45. Folloni D, Verhagen L, Mars RB, Fouragnan E, Constans C, Aubry JF, et al. Manipulation of Subcortical and Deep Cortical Activity in the Primate Brain Using Transcranial Focused Ultrasound Stimulation. Neuron. 2019;101(6):1109–16.e5.

46. Mohammadjavadi M, Ash RT, Glover GH, Pauly KB. Optimization of MR acoustic radiation force imaging (MR-ARFI) for human transcranial focused ultrasound. Magn Reson Med. 2025;94(3):1060–71.

