## Supplemental Material for "Low-Intensity Focused Ultrasound of the Amygdala in Depression and Anxiety: A First-in-Human Active-Controlled Trial"

**Table of Contents**

1. **FUS Safety protocol**
   a. Neuropsychological monitoring
2. **Blinding Considerations**
3. **Acoustic Characterization and Modeling**
   a. Acoustic Exposure Reporting (iTRUSST reporting)
   b. Acoustic Modeling
4. **Neuroimaging Methods**
   a. MRIqc
   b. fMRIPrep Preprocessing
      i. Anatomical Data
      ii. Functional Data
   c. Resting-State Connectivity Analyses
   d. Resting-State Motion Sensitivity Analysis
   e. ROI Definition
   f. Small Volume Correction Methods
5. **Individual level outcomes for arterial spin labeling**
6. **Exploratory associations between neuroimaging and self-reported symptoms of depression, anxiety, and posttraumatic stress disorder.**
7. **Supplemental Figures**
   a. **Figure S1.** Individual Trajectories of Perfusion Changes Following Low-Intensity Focused Ultrasound.
   b. **Figure S2.** Primary Auditory Cortex Activation During Target and Control Low-Intensity Focused Ultrasound.
   c. **Figure S3.** Clinical symptom and perfusion changes following amygdala-targeted FUS in a single participant.
   d. **Figure S4.** Acoustic modeling and targeting accuracy.

**1. FUS Safety Protocol**

The FUS safety protocol was designed to ensure that any unexpected observations of clinical worsening were sufficiently characterized, given the many novel elements of this study. In brief, if sustained clinically meaningful deterioration was observed, participants would cease further FUS procedures and move into this protocol, which included weekly assessments inclusive of MRI, psychiatric rating scales (including the C-SSRS), neurological examination, and neuropsychological testing (see below for further details)

First, clinically meaningful deterioration was operationally defined as worsening of >10 points on the MADRS and IDSSR, >5 points on the GAD7, or >25% change from baseline, whichever was more conservative; >2-point worsening on the CGI was also used. Per protocol, any worsening that occurred and remained at the 1-week follow up visit triggered additional monitoring. Clinically meaningful deterioration on the C-SSRS was defined as emergence of any acute suicidality, defined as “Yes” on item 4 of the C-SSRS, (i.e., active suicidal ideation with some intent to act), or any endorsement of item 5 (active ideation with specific plan and intent) or any actual, interrupted, aborted attempt or preparatory behavior; sustained worsening on the C-SSRS was not required for additional monitoring. The full IDE/FUS Safety protocol is available from the corresponding author upon reasonable request.

**1a. Neuropsychological Assessments.** Because this was a first-in-human application of FUS to this target, a multidomain neuropsychological battery was administered, loosely modeled after testing done in neurosurgical settings. The tests comprising the battery were selected in part because alternate forms were available to reduce practice effects during serial assessment of participants over five time points during enrollment (i.e., baseline, FUS to target 1 at 24h (hereafter, FUSx), FUS1 at 1-wk, FUS 2-24h, FUS2-1-wk). The test battery was as follows:

- Repeatable Battery for the Assessment of Neuropsychological Status (RBANS), Forms A, B, C, and D (1). Provides index scores of the following cognitive domains: Immediate Memory, Attention, Visuospatial/Construction, Language, and Delayed Memory and provides a Total Index score.
- Color Trails Test (CTT, parts 1 and 2), Forms A, B, C, and D (2). Assesses attention, processing speed, and cognitive flexibility.
- Mazes from the Neuropsychological Assessment Battery (NAB), Forms 1 and 2 (3). Assesses attention, processing speed, and planning.

Test form order was sequential within participant but differed across participants. For example, across the five study timepoints, “Participant A” might receive the following test order: RBANS forms B, C, D, A, B; CTT forms B, C, D, A, B; and Mazes forms 1, 2, 1, 2, 1. Whereas, “Participant B” might receive the following order: RBANS forms D, A, B, C, D; CTT forms D, A, B, C, D; and Mazes forms 2, 1, 2, 1, 2. The order of test administration remained consistent across participants, with the RBANS administered first, followed by the CTT and then the NAB Mazes.

Performance validity during testing was assessed using the RBANS Effort Index(4). A baseline score >3 was prespecified as an exclusion criterion; however, no participants met this criterion. Doctorate-level study staff were trained by the study neuropsychologists to administer and score the tests. The study neuropsychologists monitored fidelity to administration and scoring throughout the study by examining source documentation and providing written queries, instructions, and training refreshers as needed. Two neuropsychologists served as backups to the main study neuropsychologist.

A two-tiered conservative threshold was set for consideration of transitioning a participant from the main protocol to the Safety Protocol. Tier 1 was a ≥ 0.5 standard deviation decline from Baseline on any of the RBANS cognitive domain index or total scores (see above) or CTT. Tier 2 was that this decline had to occur both at 24 hours post FUS1 or FUS2 and sustained in the same RBANS domain score or Maze at 1 week post FUS1 or FUS2. Hitting this 2-tier threshold triggered an *a priori* in-depth algorithmic review by the main study neuropsychologist to ensure that the change was not due to performance invalidity, documented extraneous factors (e.g., environmental disruption), and determination that the 0.5 SD decline met or exceeded the critical values required for statistical significance between RBANS alternate forms (5). The algorithm also requires review of all RBANS subtest scores and CTT scores for possible worrisome signs determined by expert clinical judgment and not reflected in the RBANS domain and total scores. Based on the results of the algorithmic review, the main study neuropsychologist rendered a decision to either continue the participant in the standard FUS protocol or transfer the participant to the FUS Safety Protocol. In either event, another study neuropsychologists independently reviewed the main neuropsychologists’ opinion and rationale and rendered and “agree or not agree” opinion. These reviews were sent to the Principal Investigator who rendered the final decision.

**2. Blinding Considerations**

As detailed in the manuscript, this proposal was designed to answer the question of precision prior to efficacy. To this end, we utilized an active-controlled design, comparing effect of FUS on the target (right amygdala) compared to a cortical control (S1). We chose contralateral S1 for our control region because of its distance to the amygdala, lack of involvement in FMRI paradigms, and its more modest contribution to core neurobiology of depression (compared to the amygdala). We recognize this complicated blinding as it requires placing the transducer on different locations on the head. We managed this through several elements; first, primary outcome measures were blinded (MADRS for depression; GAD7 and PCL-5 were both self-rated). When presenting the study to participants, we sought to manage expectations by telling them we were looking for which region worked the best and we were trying two different approaches; all participants were informed they were receiving active (verum) FUS. We did not have participants fill out a blinding inquiry since no sham was used. Based on our experience, it is possible to imagine that future FUS studies may require the use of an active control and a sham.

**3. Acoustic Characterization and Modeling**

**3a. Acoustic Exposure Reporting (iTRUSST)**

FUS was delivered using the Brainsonix BX1002/Pulsar 1002 system (Brainsonix, Los Angeles, CA) with a circular, single-element, spherically focused transducer manufactured by Blatek (State College, PA) (6). The transducer had a 61-mm active aperture and a nominal focal length/radius of curvature of 65 mm. The Brainsonix platform incorporates an integrated ultrasonic drive generator with programmable carrier frequency, pulse width, pulse-repetition frequency, and drive amplitude. The system calculates the required electrical drive voltage from the prescribed acoustic exposure parameters using transducer-specific calibration data rather than requiring manual voltage selection.

The drive generator produces a bipolar, pulsed quasi-sinusoidal waveform. The core timing and control electronics use a programmable logic device, with carrier-frequency generation provided by direct digital synthesis. The system incorporates hardware and software monitoring of drive voltage and output parameters and disables transmission if predefined limits are exceeded.

Each stimulator, transducer, cable, and MRI filter configuration was individually calibrated. During calibration, a calibrated hydrophone was used to identify the spatial location of maximum acoustic output, characterize acoustic pressure and intensity at that location, and map the −6 dB spatial extent of the ultrasound field. The Brainsonix calibration procedure uses PVDF bilaminar membrane hydrophones for focal-zone characterization and voltage-pressure measurements at lower drive levels, with a scattering hydrophone used at higher output levels after cross-calibration against the bilaminar hydrophone. Cable and MRI wall-filter configurations are incorporated into the calibration because these components can materially affect focal acoustic output.

Independent free-field acoustic characterization was performed by Acertara Acoustic Laboratories, an ISO 17025-accredited laboratory. For the 65-mm nominal-focus Brainsonix probe, measurements were obtained using a calibrated hydrophone (S/N S5-186; sensitivity −266.94 dB re 1 µV/Pa). The measured center frequency was 0.650 MHz with a −3 dB bandwidth of 0.019 MHz. The study-specific free-field measurements demonstrated a focal peak depth of 60.0 mm, with axial −6 dB boundaries at 46.9 and 77.0 mm, corresponding to an axial −6 dB focal length of 30.1 mm. The transverse −6 dB focal width was 4.0 mm. These values were used to characterize the expected focal region for treatment planning.

For the study protocol, the transducer operated at a carrier frequency of 650 kHz, PRF of 10 Hz, pulse width of 5 ms, and 5% duty cycle. Sonication was delivered in 30-second on/30-second off blocks, with two 10-minute applications per session. Device output was set to a free-field spatial-peak temporal-average intensity of 937.9 mW/cm² and spatial-peak pulse-average intensity of 18.84 W/cm². Using the conventional 0.3 dB/cm/MHz derating approach, these corresponded to ISPTA.3=719.5 mW/cm² and ISPPA.3=14.39 W/cm², with a derated peak negative pressure of 0.61 MPa and mechanical index of 0.76. These are the same exposure parameters reported in the primary Methods. Thermal profiles were individually generated in Babelbrain and can be provided upon request.

**3b. Acoustic modeling considerations**

This study used post-hoc acoustic modeling; this was done for several reasons. Primarily, at the time of study inception (2021), acoustic modeling was not yet standardized or readily available. While the emergence of programs (e.g., Babelbrain, K-Plan, M-Sound) have since been developed and released, these were not available at the time. We felt that a substantial change to the methods during the conduct of the study would be unwarranted.

**4. Neuroimaging Methods**

MRIqc

All collected research MRI data have been run through MRIqc, a fully automated pipeline designed to assess data quality and enable visual examination of MRI scans. MRIqc provides a standardized, algorithmic and subjective approach to quality assurance of MRI data. The pipeline collects information and extracts Image Quality Metrics (IQMs) grouped in four broad categories: noise, information theory and spatial distribution, specific artifacts, and additional measures such as tissue distributions. Visual reports that are generated for each subject allow us to examine potential quality issues.  IQMs are extracted at a subject-level, then group-level reports are generated. The group-level visual reports provide box-and-whisker plots of each IQM to show the distribution of subjects for each IQM, then we can use these plots to identify subjects that may be outliers.

Overall, data quality across acquisition conditions was acceptable. Signal metrics (tSNR and SNR) were within expected ranges, and motion was low across the majority of participants. Observed variability was driven by a small number of identifiable outliers. One subject was flagged for deviations in signal and/or motion metrics but, following visual inspection, was retained because no significant artifacts, preprocessing failures, or other data quality concerns were identified.

fMRI Preprocessing

Results included in this manuscript come from preprocessing performed using *fMRIPrep* 24.1.1 (7) which is based on *Nipype* 1.8.6 (8).

*Anatomical data preprocessing*

Each T1w image was corrected for intensity non-uniformity (INU) with N4BiasFieldCorrection (9), distributed with ANTs 2.5.3 (10) . The T1w-reference was then skull-stripped with a *Nipype* nimplementation of the antsBrainExtraction.sh workflow (from ANTs), using OASIS30ANTs as target template. Brain tissue segmentation of cerebrospinal fluid (CSF), white-matter (WM) and gray-matter (GM) was performed on the brain-extracted T1w using fast (11) (FSL) . An anatomical T1w-reference map was computed after registration of 3 <module ‘nipype.interfaces.image’ from ‘/opt/conda/envs/fmriprep/lib/python3.11/site-packages/nipype/interfaces/image.py’> images (after INU-correction) using mri_robust_template (12) (FreeSurfer 7.3.2). Brain surfaces were reconstructed using recon-all (13) (FreeSurfer 7.3.2), and the brain mask estimated previously was refined with a custom variation of the method to reconcile ANTs-derived and FreeSurfer-derived segmentations of the cortical gray-matter of Mindboggle (14). Volume-based spatial normalization to one standard space (MNI152NLin2009cAsym) was performed through nonlinear registration with antsRegistration (ANTs 2.5.3), using brain-extracted versions of both T1w reference and the T1w template. The following template was selected for spatial normalization and accessed with *TemplateFlow* (15): *ICBM 152 Nonlinear Asymmetrical template version 2009c* (16).

*Functional Data preprocessing*

The following preprocessing was performed for functional MRI data. First, a reference volume was generated, using a custom methodology of *fMRIPrep*, for use in head motion correction. Head-motion parameters with respect to the BOLD reference (transformation matrices, and six corresponding rotation and translation parameters) are estimated before any spatiotemporal filtering using mcflirt (17). The BOLD reference was then co-registered to the T1w reference using bbregister (FreeSurfer) which implements boundary-based registration (18). Co-registration was configured with six degrees of freedom. Several confounding time-series were calculated based on the *preprocessed BOLD*: framewise displacement (FD), DVARS and three region-wise global signals. FD was computed using two formulations following Power (absolute sum of relative motions, (19)) and Jenkinson (relative root mean square displacement between affines, (17) FD and DVARS are calculated for each functional run, both using their implementations in *Nipype*. The three global signals are extracted within the CSF, the WM, and the whole-brain masks. Additionally, a set of physiological regressors were extracted to allow for component-based noise correction (*CompCor*,(20)). Principal components are estimated after high-pass filtering the *preprocessed BOLD*time-series (using a discrete cosine filter with 128s cut-off) for the two *CompCor* variants: temporal (tCompCor) and anatomical (aCompCor). tCompCor components are then calculated from the top 2% variable voxels within the brain mask. For aCompCor, three probabilistic masks (CSF, WM and combined CSF+WM) are generated in anatomical space. The implementation differs from that of Behzadi et al. in that instead of eroding the masks by 2 pixels on BOLD space, a mask of pixels that likely contain a volume fraction of GM is subtracted from the aCompCor masks. This mask is obtained by dilating a GM mask extracted from the FreeSurfer’s *aseg* segmentation, and it ensures components are not extracted from voxels containing a minimal fraction of GM. Finally, these masks are resampled into BOLD space and binarized by thresholding at 0.99 (as in the original implementation). Components are also calculated separately within the WM and CSF masks. For each CompCor decomposition, the *k* components with the largest singular values are retained, such that the retained components’ time series are sufficient to explain 50 percent of variance across the nuisance mask (CSF, WM, combined, or temporal). The remaining components are dropped from consideration. The head-motion estimates calculated in the correction step were also placed within the corresponding confounds file. The confound time series derived from head motion estimates and global signals were expanded with the inclusion of temporal derivatives and quadratic terms for each (21). Frames that exceeded a threshold of 0.5 mm FD or 1.5 standardized DVARS were annotated as motion outliers. Additional nuisance timeseries are calculated by means of principal components analysis of the signal found within a thin band (*crown*) of voxels around the edge of the brain, as proposed by (22). All resamplings can be performed with *a single interpolation step* by composing all the pertinent transformations (i.e. head-motion transform matrices, susceptibility distortion correction when available, and co-registrations to anatomical and output spaces). Gridded (volumetric) resamplings were performed using nitransforms, configured with cubic B-spline interpolation.

Resting-State Functional Connectivity Analyses using CONN

Analyses of fMRI data were performed using CONN (23) release 22.v2407 and SPM release 25.25.01.rc3.

Preprocessing: Functional data were smoothed using spatial convolution with a Gaussian kernel of 5 mm full width half maximum (FWHM).

Denoising: In addition, functional data were denoised using a standard denoising pipeline including the regression of potential confounding effects characterized by white matter timeseries (5 CompCor noise components), CSF timeseries (5 CompCor noise components), realign regressors and their first order derivatives (12 components), session and task effects and their first order derivatives (6 factors), and linear trends (2 factors) within each functional run, followed by bandpass frequency filtering of the BOLD timeseries (24) between 0.008 Hz and 0.09 Hz. CompCor (20, 25) noise components within white matter and CSF were estimated by computing the average BOLD signal as well as the largest principal components orthogonal to the BOLD average within each subject's eroded segmentation masks. From the number of noise terms included in this denoising strategy, the effective degrees of freedom of the BOLD signal after denoising were estimated to range from 227.3 to 341 (average 324.5) across all subjects.

First-level analysis SBC_01: Seed-based connectivity maps (SBC) and ROI-to-ROI connectivity matrices (RRC) were estimated characterizing the patterns of functional connectivity with 10 ROIs. Functional connectivity strength was represented by Fisher-transformed bivariate correlation coefficients from a weighted general linear model (weighted-GLM), defined separately for each pair of seed and target areas, modeling the association between their BOLD signal timeseries. Individual scans were weighted by a boxcar signal characterizing each individual task or experimental condition convolved with an SPM canonical hemodynamic response function and rectified.

Group-level analyses were performed using a General Linear Model (GLM). For each individual voxel a separate GLM was estimated, with first-level connectivity measures at this voxel as dependent variables (one independent sample per subject and one measurement per task or experimental condition, if applicable), and groups or other subject-level identifiers as independent variables. Voxel-level hypotheses were evaluated using multivariate parametric statistics with random-effects across subjects and sample covariance estimation across multiple measurements. Inferences were performed at the level of individual clusters (groups of contiguous voxels). Cluster-level inferences were based on parametric statistics from Gaussian Random Field theory (26). Results were thresholded using a combination of a cluster-forming p < 0.001 voxel-level threshold, and a familywise corrected p-FDR < 0.05 cluster-size threshold (27).

Resting-State Motion Sensitivity Analysis

Mean framewise displacement (FD) was compared between the baseline and post-amygdala FUS resting-state scans using a paired-samples *t*-test (N = 10). Mean FD was 0.147 mm (SD = 0.054) at baseline and 0.176 mm (SD = 0.073) following amygdala FUS, representing a mean within-subject increase of 0.029 mm (SD = 0.064). The paired-samples *t*-test did not indicate a significant difference in FD between sessions, *t*(9) = 1.43, *p* = .187. A Wilcoxon signed-rank test yielded a consistent result (*V* = 18.0, *p* = .375). Thus, there was no evidence of a systematic increase in head motion following amygdala FUS.

Because the primary resting-state connectivity findings involved sensorimotor regions, we additionally conducted a sensitivity analysis to assess whether between-session differences in head motion could account for the observed effects. Mean framewise displacement (FD) was calculated for each resting-state scan, and the change in FD between sessions was included as a second-level covariate in the seed-to-voxel analysis. Using the same cluster-level threshold as the primary analysis (*p* < .05, FDR-corrected), but a more lenient voxelwise threshold of *p* < .005 rather than *p* < .001, the observed decreases in connectivity between the basolateral amygdala and the supplementary motor area (SMA), precentral gyrus, and postcentral gyrus remained significant after accounting for between-session changes in FD. The SMA finding was observed at the same peak coordinate as in the primary analysis, while the peak coordinates for the precentral and postcentral gyrus findings differed slightly but remained within the corresponding anatomical regions.

Region of interest (ROI) Definition

ROIs were defined a priori or based on independent anatomical or functional references, as specified below.

- **Amygdala (ASL functional ROI):** The amygdala ROI was functionally defined for each participant using the individual-level ASL contrast comparing amygdala FUS with baseline (amygdala FUS > baseline). A basolateral amygdala (BLA) mask (described below) was used to constrain the search region. The ASL maps were visually inspected within the BLA mask, and voxels showing apparent activation following FUS were identified to define the participant-specific functional ROI. No fixed statistical or cluster-size threshold was applied because the extent and magnitude of ASL activation within the BLA varied substantially across participants. Consequently, the spatial extent of the functional ROI varied across participants.
- **Hippocampal head:** The hippocampal head ROI was defined anatomically as previously described (28).
- **Basolateral amygdala (BLA):** The BLA ROI was defined using the Juelich Histological Atlas (29, 30) and thresholded at 50% probability.
- **Primary somatosensory cortex (S1):** The S1 ROI was defined as a 10-mm-radius sphere centered on the peak coordinate (x=-52, y=-22, z =44) identified using Neurosynth.
- **Insula:** The insula ROI was defined using the Harvard-Oxford cortical atlas.
- **Rostral anterior cingulate cortex (rACC):** The rACC ROI was defined as a 5-mm-radius sphere centered on the peak rACC coordinate (x=12, y=36, z=8) identified from the group-level ASL analysis.
- **Ventromedial prefrontal cortex (vmPFC):** The vmPFC ROI was defined using the functional parcellation of medial frontal cortex described by de la Vega et al. (2016). Specifically, we used the **k = 9** solution, in which *k* denotes the number of functionally distinct clusters identified through k-means clustering of whole-brain coactivation patterns. This ROI corresponds to cluster 8 of the solution.
- **Primary auditory cortex:** Bilateral primary auditory cortex was defined using the Juelich Histological Atlas(29, 30) and thresholded at 50% probability.

Small-Volume Correction

Small-volume correction (SVC) was conducted separately for each analysis using the defined ROI masks described above. The number and selection of SVC masks differed across analyses based on the study's a priori hypotheses and follow-up of findings from uncorrected whole-brain analyses.

For ASL group-level analyses, SVC was performed separately within seven ROI masks: bilateral amygdala, bilateral thalamus, right rostral anterior cingulate cortex (rACC), right ventromedial prefrontal cortex (vmPFC), and left putamen. Bilateral amygdala and right vmPFC were specified a priori. Bilateral thalamus, right rACC, and left putamen were included as exploratory follow-up ROIs based on activation observed in the uncorrected whole-brain analysis. SVC was performed using FSL Randomise within each ROI, with statistical significance defined as family-wise error (FWE)-corrected p < .05.

For the FUS on vs. FUS off analysis, SVC was performed separately within five ROI masks: left amygdala, left thalamus, right vmPFC, right rACC, and left insula. Left amygdala and right vmPFC were specified a priori. Left thalamus, right rACC, and left insula were included as exploratory follow-up ROIs based on activation observed in the uncorrected whole-brain analysis. SVC was performed in SPM25 using the small-volume correction procedure, with statistical significance defined as FWE-corrected p < .05.

**5. Individual level ASL findings**

We observed significant individual variability in terms of the direction of ASL findings. For example, approximately half of the participants demonstrated a reduction in ASL signal when compared to their baseline; whereas the other demonstrated an increase (Figure S1A**)**. Because the net magnitude of change was greater in the positive direction, the group-level findings appeared to be an increase in ASL, yet individual directions indicate a more complicated and relationship between beam and perfusion response. This was in contrast when evaluating ASL signal in the active control (S1); in that condition, did not observe any changes in, at any stage of participation (**Figure S1B**). In both figures, note that half of participants received amygdala to S1 and the other S1 to amygdala.

**6. Exploratory associations between neuroimaging and self-reported symptoms of depression, anxiety, and posttraumatic stress disorder.**

As an exploratory analysis, we examined associations between changes in neuroimaging measures following amygdala sonication and clinical measures assessed at 24 hours and 1-week post-sonication. Changes in amygdala perfusion reflected the magnitude of change from baseline following sonication. Changes in resting-state functional connectivity (rsFC) were calculated as post-sonication minus baseline, with negative values reflecting decreases in connectivity. Clinical symptom change scores were calculated as post-sonication minus baseline, with negative values reflecting reductions in symptom severity. The Clinical Global Impression–Improvement (CGI-I) was analyzed as a post-sonication improvement rating and was not calculated as a change score.

**Amygdala perfusion.** Changes in amygdala perfusion were significantly associated with changes in CGI-S ratings at 24 hours (r=.773, p=.009) and 1-week post-sonication (r=.664, p=.036). Changes in amygdala perfusion were significantly associated with CGI-I ratings at 24 hours post amygdala-targeted FUS (*r* = .904, *p* < .001) but not at 1-week (r=.417, p=.230). Associations with changes in GAD-7 scores were observed; this was trend level at 24 hours (*r* = .592, *p* = .071) and significant at 1 week (*r* = .648, *p* = .043). Changes in amygdala perfusion were significantly associated with changes in PCL-5 scores at 24 hours (*r* = .853, *p* = .002), with the association at 1 week reaching trend level (*r* = .610, *p* = .061). Changes in amygdala perfusion were also significantly associated with changes IDSSR scores at 24 hours (*r* = .806, *p* = .005) and 1 week (*r* = .794, *p* = .006).

**Resting-state functional connectivity.** Changes in rsFC were characterized by decreases in connectivity following amygdala sonication relative to baseline. The decrease in connectivity between the right basolateral amygdala (BLA) and right precentral gyrus was associated with CGI-I ratings at 24 hours at trend level (*r* = .603, *p* = .065). No significant associations were observed between changes in right BLA–right precentral gyrus rsFC and the other clinical measures at either time point. Similarly, no significant associations were observed between changes in right BLA–left supplementary motor area (SMA) rsFC and clinical measures. Changes in right BLA–left postcentral gyrus rsFC were significantly associated with MADRS scores at 24 hours, as reported in the main manuscript, but were not significantly associated with any other clinical measures.

**8. Supplementary Figures**

S1. Individual Trajectories of Perfusion Changes Following Low-Intensity Focused Ultrasound.

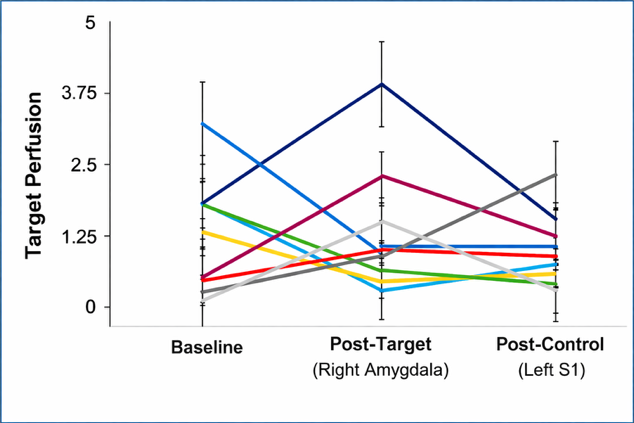

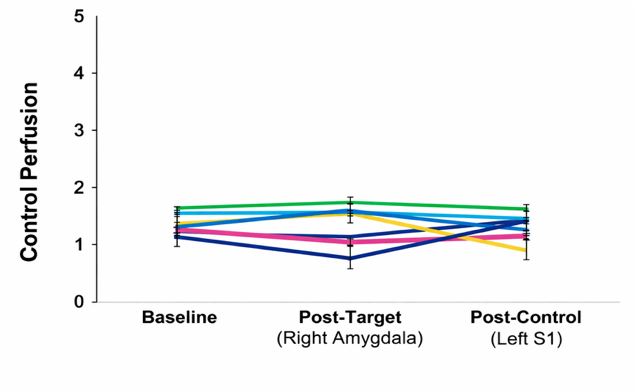

A

B

Key: S1, Primary somatosensory cortex.

S1 Figure Legend. (A) Individual trajectories of ASL signal in the target location (right amygdala) at baseline, post target (right amygdala) sonication, and post control (left S1) sonication. (B) Individual trajectories of ASL signal in the control location (left S1) at baseline, post target (right amygdala) sonication, and post control (left S1) sonication.

S2. Primary Auditory Cortex Activation During Target and Control Low-Intensity Focused Ultrasound.

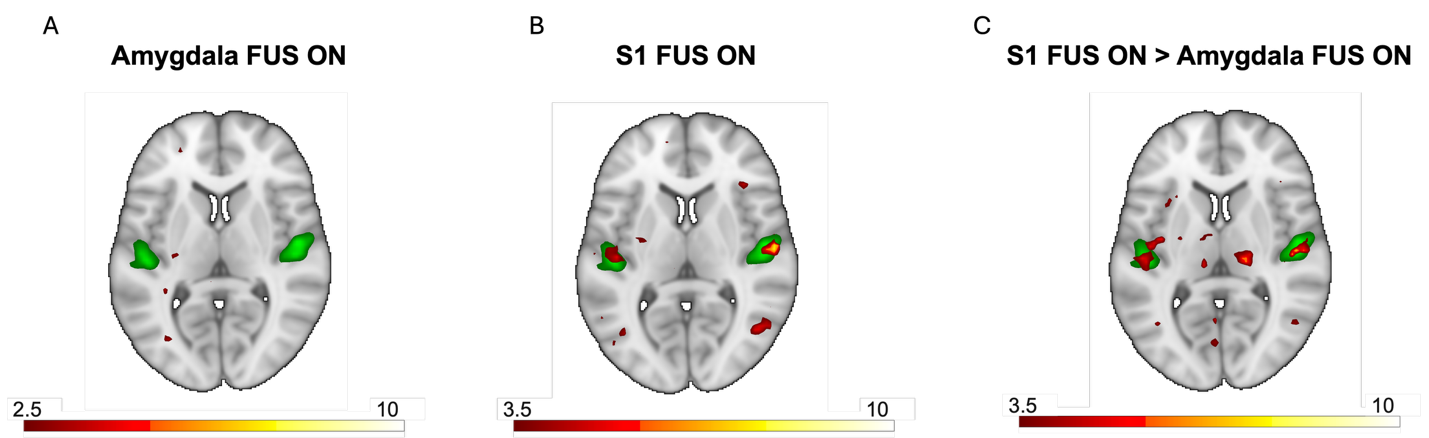

Key: FUS, low intensity focused; S1 primary somatosensory cortex.

S2 Figure Legend. Green represents the primary auditory cortex region of interest. (A) No significant activation of the primary auditory cortex was observed during target FUS to the right amygdala. (B) Significant activation of the primary auditory cortex was observed during control FUS to the left somatosensory cortex (S1). (C) Activation of the primary auditory cortex was significantly greater during control FUS than target FUS.

S3. Clinical symptom and perfusion changes following amygdala-targeted FUS in a single participant.

B

A

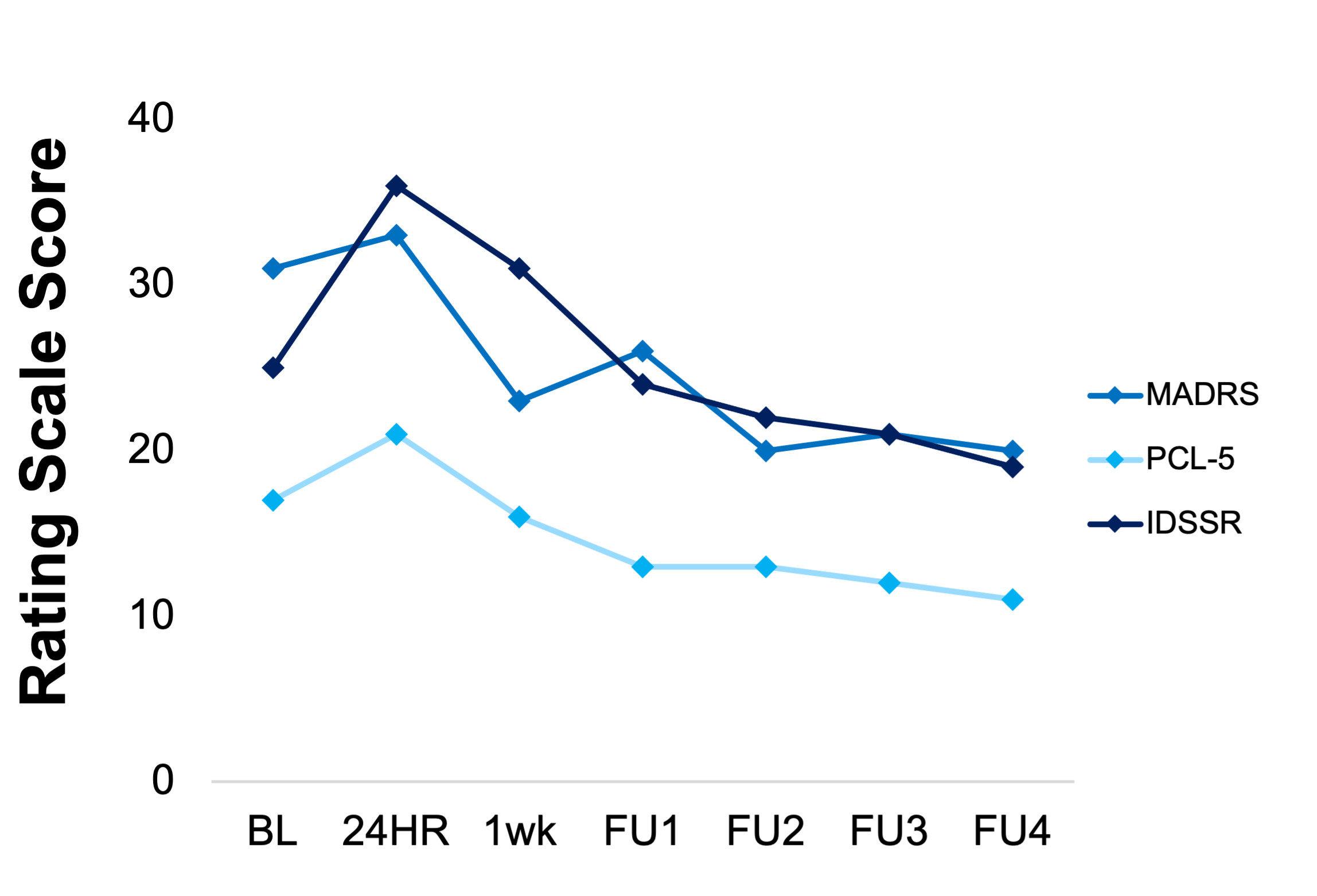

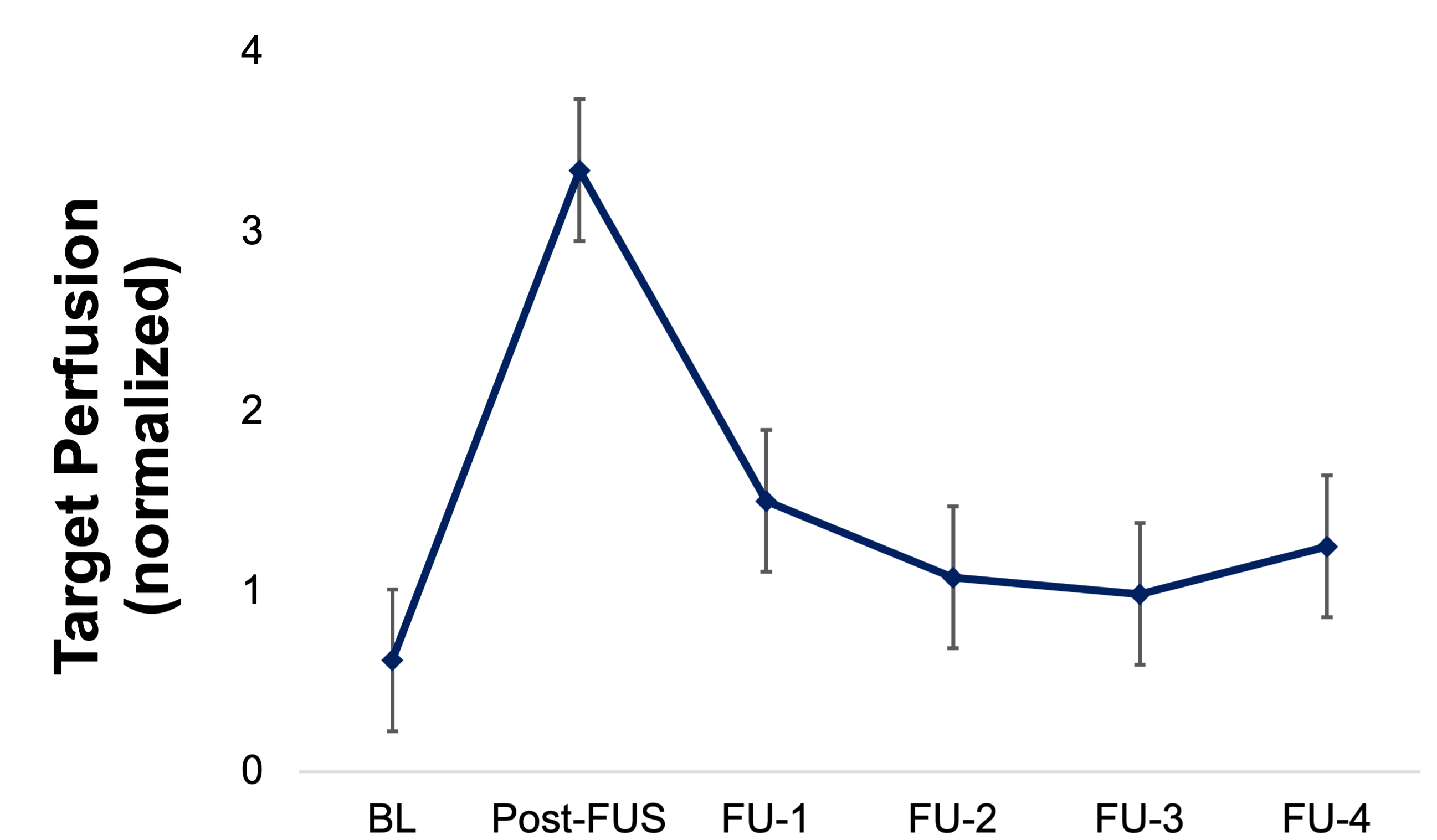

Key: MADRS, Montgomery-Åsberg Depression Rating Scale; PCL-5, PTSD Checklist for DSM-5; IDSSR, inventory of depressive symptomatology, self-report; BL, baseline; FU, follow-up; FUS, low intensity focused ultrasound.

S3 Figure Legend. (A) MADRS, PCL, and IDSSR scores at baseline, 24 hours and 1 week following amygdala-targeted FUS, and at four subsequent extended follow-up assessments. The most pronounced increase was observed on the IDSSR at 24 hours and 1 week following FUS, with scores returning toward baseline during the extended follow-up period. (B) Perfusion in the right amygdala at baseline, following amygdala-targeted FUS, and at four subsequent extended follow-up assessments. Perfusion increased following FUS and returned toward baseline during the extended follow-up period.

S4. Acoustic modeling and targeting accuracy.

B

A

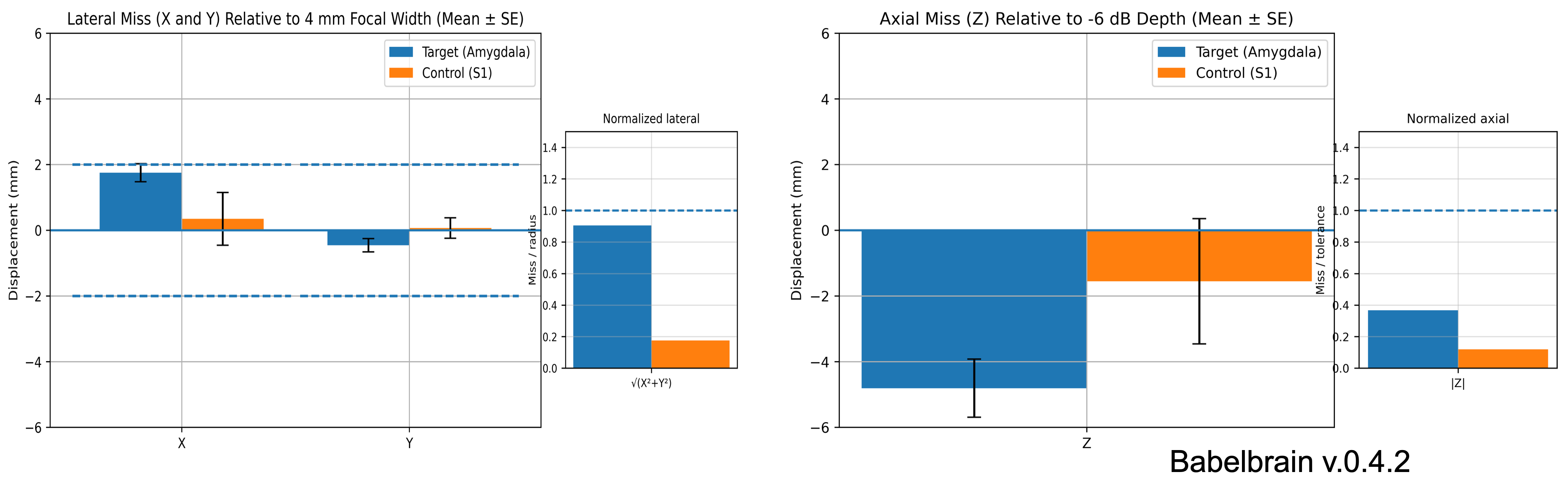

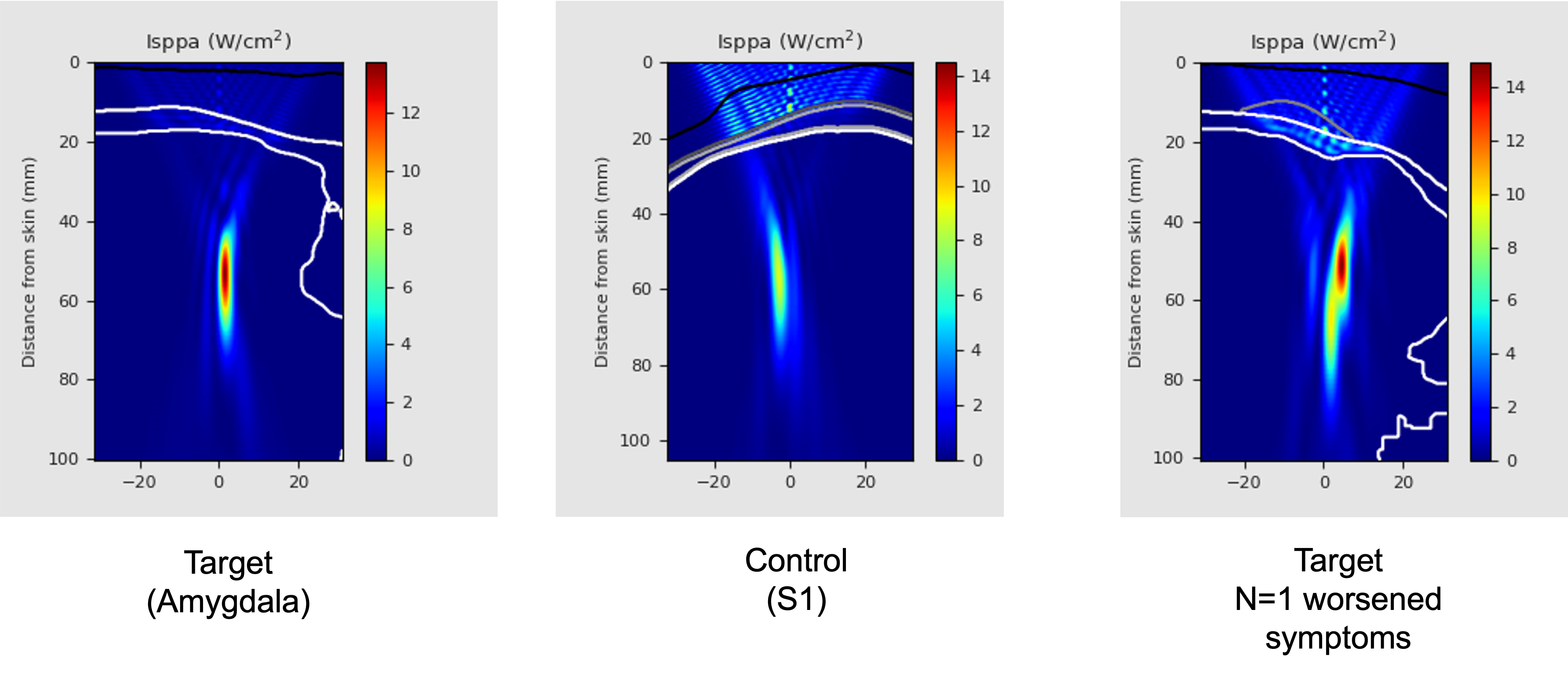

Key: S1, primary somatosensory cortex; Isppa, Spatial-Peak Pulse-Average Intensity

S4 Figure Legend. (A) Displacement between line-of-sight target locations and modeled beam locations across participants in the x-, y-, and z-directions. B) Representative modeled beam locations for the target and control conditions, alongside the modeled beam location for the target condition in the participant who demonstrated clinically significant worsening in depressive symptoms following amygdala-targeted FUS.

Supplemental References

1.	Randolph C, Tierney MC, Mohr E, Chase TN. The Repeatable Battery for the Assessment of Neuropsychological Status (RBANS): preliminary clinical validity. J Clin Exp Neuropsychol. 1998;20(3):310-9.

2.	D’Elia LF, Satz, P., Uchiyama, C.L., & White, T. . Color Trails Test. Psychological Assessment Resources. 1996.

3.	T SRW. NAB Neuropsychological Assessment Battery: Administration, scoring, and Interpretation Manual. Psychological Assessment Resources. 2003.

4.	Silverberg ND, Wertheimer JC, Fichtenberg NL. An effort index for the Repeatable Battery For The Assessment Of Neuropsychological Status (RBANS). Clin Neuropsychol. 2007;21(5):841-54.

5.	Randolph C. RBANS Update: The Repeatable Battery for the Assessment of Neuropsychological Status. Pearson. 2012.

6.	Schafer ME, Spivak NM, Korb AS, Bystritsky A. Design, Development, and Operation of a Low-Intensity Focused Ultrasound Pulsation (LIFUP) System for Clinical Use. IEEE Trans Ultrason Ferroelectr Freq Control. 2021;68(1):54-64.

7.	Esteban O, Markiewicz CJ, Blair RW, Moodie CA, Isik AI, Erramuzpe A, et al. fMRIPrep: a robust preprocessing pipeline for functional MRI. Nat Methods. 2019;16(1):111-6.

8.	Gorgolewski K, Burns CD, Madison C, Clark D, Halchenko YO, Waskom ML, et al. Nipype: a flexible, lightweight and extensible neuroimaging data processing framework in python. Front Neuroinform. 2011;5:13.

9.	Tustison NJ, Avants BB, Cook PA, Zheng Y, Egan A, Yushkevich PA, et al. N4ITK: improved N3 bias correction. IEEE Trans Med Imaging. 2010;29(6):1310-20.

10.	Avants BB, Epstein CL, Grossman M, Gee JC. Symmetric diffeomorphic image registration with cross-correlation: evaluating automated labeling of elderly and neurodegenerative brain. Med Image Anal. 2008;12(1):26-41.

11.	Zhang Y, Brady M, Smith S. Segmentation of brain MR images through a hidden Markov random field model and the expectation-maximization algorithm. IEEE Trans Med Imaging. 2001;20(1):45-57.

12.	Reuter M, Rosas HD, Fischl B. Highly accurate inverse consistent registration: a robust approach. Neuroimage. 2010;53(4):1181-96.

13.	Dale AM, Fischl B, Sereno MI. Cortical surface-based analysis. I. Segmentation and surface reconstruction. Neuroimage. 1999;9(2):179-94.

14.	Klein A, Ghosh SS, Bao FS, Giard J, Hame Y, Stavsky E, et al. Mindboggling morphometry of human brains. PLoS Comput Biol. 2017;13(2):e1005350.

15.	Ciric R, Thompson WH, Lorenz R, Goncalves M, MacNicol EE, Markiewicz CJ, et al. TemplateFlow: FAIR-sharing of multi-scale, multi-species brain models. Nat Methods. 2022;19(12):1568-71.

16.	Fonov V, Evans AC, Botteron K, Almli CR, McKinstry RC, Collins DL, et al. Unbiased average age-appropriate atlases for pediatric studies. Neuroimage. 2011;54(1):313-27.

17.	Jenkinson M, Bannister P, Brady M, Smith S. Improved optimization for the robust and accurate linear registration and motion correction of brain images. Neuroimage. 2002;17(2):825-41.

18.	Greve DN, Fischl B. Accurate and robust brain image alignment using boundary-based registration. Neuroimage. 2009;48(1):63-72.

19.	Power JD, Mitra A, Laumann TO, Snyder AZ, Schlaggar BL, Petersen SE. Methods to detect, characterize, and remove motion artifact in resting state fMRI. Neuroimage. 2014;84:320-41.

20.	Behzadi Y, Restom K, Liau J, Liu TT. A component based noise correction method (CompCor) for BOLD and perfusion based fMRI. Neuroimage. 2007;37(1):90-101.

21.	Satterthwaite TD, Elliott MA, Gerraty RT, Ruparel K, Loughead J, Calkins ME, et al. An improved framework for confound regression and filtering for control of motion artifact in the preprocessing of resting-state functional connectivity data. Neuroimage. 2013;64:240-56.

22.	Patriat R, Reynolds RC, Birn RM. An improved model of motion-related signal changes in fMRI. Neuroimage. 2017;144(Pt A):74-82.

23.	Whitfield-Gabrieli S, Nieto-Castanon A. Conn: a functional connectivity toolbox for correlated and anticorrelated brain networks. Brain Connect. 2012;2(3):125-41.

24.	Hallquist MN, Hwang K, Luna B. The nuisance of nuisance regression: spectral misspecification in a common approach to resting-state fMRI preprocessing reintroduces noise and obscures functional connectivity. Neuroimage. 2013;82:208-25.

25.	Chai XJ, Castanon AN, Ongur D, Whitfield-Gabrieli S. Anticorrelations in resting state networks without global signal regression. Neuroimage. 2012;59(2):1420-8.

26.	Worsley KJ, Marrett S, Neelin P, Vandal AC, Friston KJ, Evans AC. A unified statistical approach for determining significant signals in images of cerebral activation. Hum Brain Mapp. 1996;4(1):58-73.

27.	Chumbley J, Worsley K, Flandin G, Friston K. Topological FDR for neuroimaging. Neuroimage. 2010;49(4):3057-64.

28.	Ritchey M, Montchal ME, Yonelinas AP, Ranganath C. Delay-dependent contributions of medial temporal lobe regions to episodic memory retrieval. Elife. 2015;4.

29.	Eickhoff SB, Paus T, Caspers S, Grosbras MH, Evans AC, Zilles K, et al. Assignment of functional activations to probabilistic cytoarchitectonic areas revisited. Neuroimage. 2007;36(3):511-21.

30.	Eickhoff SB, Stephan KE, Mohlberg H, Grefkes C, Fink GR, Amunts K, et al. A new SPM toolbox for combining probabilistic cytoarchitectonic maps and functional imaging data. Neuroimage. 2005;25(4):1325-35.
